# Early-life critical windows of susceptibility to metal mixture exposure and neurocognitive mechanisms underlying adolescent risky decision-making

**DOI:** 10.64898/2026.09.12.26362916

**Authors:** Kristie Oluyemi, Erik de Water, Elza Rechtman, Azzurra Invernizzi, Vida Rebello, Michelle A. Rodriguez, Libni A. Torres-Olascoaga, Luis Bautista-Arredondo, Sandra Martínez-Medina, Rafael Lara-Estrada, Erika Proal, Viviana Villicaña-Muñoz, Chris Gennings, Cheuk Y. Tang, Daniela Schiller, Abraham Reichenberg, Roberta F. White, Francisco X. Castellanos, Martha M. Téllez-Rojo, Robert O. Wright, Manish Arora, Megan K. Horton

## Abstract

**Introduction:** The developing neural circuitry supporting adolescent risk-taking is vulnerable to metals, yet few studies have examined how early-life metal mixture exposure affects neural mechanisms underlying risk-taking. We used novel dentine biomarkers to identify critical windows of early-life metal mixture exposure associated with neurocognitive correlates of risky decision-making in early adolescence.

**Methods:** Participants (ages 8–14, 45% female) were recruited from the Programming Research in Obesity, Growth, Environment and Social Stressors (PROGRESS) longitudinal birth cohort in Mexico City. Weekly concentrations of barium (Ba), copper (Cu), lithium (Li), magnesium (Mg), manganese (Mn), lead (Pb), tin (Sn), strontium (Sr), and zinc (Zn) were measured in children’s deciduous teeth from 19 weeks pre-birth to 43 weeks postnatally. Neural activity during risky decision-making and reward processing was assessed using a functional magnetic resonance imaging gambling paradigm. Risk-taking tendency, risk sensitivity, and reward sensitivity were estimated from participants’ decisions. We used lagged weighted quantile sum regression to examine time-varying associations between metal mixture exposure and neurocognitive outcomes, adjusting for child age, child sex, and socioeconomic status.

**Results:** Higher metal mixture exposure during the early to late postnatal period was associated with increased risk-taking tendency and risk sensitivity. Exposure during the prenatal and late postnatal periods was associated with increased reward sensitivity and reduced activation in the angular gyrus/inferior parietal lobule during risky decision-making. Exposure during the late postnatal period was associated with increased activation in the insula/rolandic operculum during reward processing. Postnatal associations for risk-related and reward-related neural activation were primarily driven by Sr and Mg respectively, while postnatal associations for risk-taking tendency, risk sensitivity, and reward sensitivity were driven by multiple metals in the mixture.

**Conclusions:** Early-life metal mixture exposure is differentially associated with neurocognitive correlates of risky decision-making in adolescence, depending on exposure timing.

## Introduction

Heightened risk-taking during adolescence is a normative phenomenon supporting the progression towards self-sufficiency and independence critical for adulthood.^1,2^ While risk-taking in adolescence is thought to be evolutionarily adaptive, maladaptive risk-taking, defined as risk-taking behaviors leading to adverse health outcomes (e.g., substance abuse, reckless driving), accounts for the majority of adolescent fatalities among developed countries,^3–7^ and increases the risk for poorer later life outcomes, including reduced educational attainment and economic instability.^7–10^ Several studies have observed similar developmental patterns of risk-taking across cultures, suggesting that biological factors such as brain maturational trajectories may contribute to this phenomenon. ^6,11,12^ However, as prior research has linked early-life exposure to essential and non-essential metals with structural and functional brain changes in childhood and adolescence,^13–16^ susceptibility to maladaptive risk-taking may also be influenced by environmentally-induced neurodevelopmental changes. The Developmental Origins of Health and Disease (DOHaD) hypothesis posits that early-life environmental insults can disrupt biological developmental pathways with lasting effects into later life.^17,18^ Given the rapid structural and physiological brain changes from gestation to adolescence,^19–23^ early-life metal exposures may represent an important, understudied risk factor conferring vulnerability to maladaptive adolescent risk-taking. Further, as the most substantial brain development occurs during the first two years of life,^24^ the prenatal and infancy periods are particularly sensitive to metal toxicants and may represent critical periods of susceptibility.^25^ Examining adolescent risk-taking from a DoHaD perspective will therefore advance understanding of how early-life environmental factors shape and adolescent brain and behavior, and inform prevention strategies to mitigate the development of maladaptive risk-taking phenotypes and risk-taking related psychopathology.

The emergence of heightened risk-taking behavior during adolescence is often attributed to a developmental imbalance between early-maturing brain reward motivational systems (e.g., ventral striatum and medial prefrontal cortex) and relatively later maturing cognitive control systems (e.g., lateral prefrontal cortex). ^3,26,27^ However, this imbalance is not observed in all adolescents.^28^ Only a subset of youth engages in maladaptive risk-taking behaviors,^1,29–32^ suggesting that individual differences in early-life environmental factors may shape vulnerability to maladaptive adolescent risk-taking. Early-life exposure to neurotoxic metals such as lead (Pb) and manganese (Mn) has been associated with cognitive and behavioral factors implicated in adolescent risk-taking, including increased externalizing symptoms (e.g., rule-breaking, impulsivity),^33–37^ increased reward motivation,^38^ and reduced inhibitory control.^39–43^ Neuroimaging studies have also observed associations between early-life metal exposure and alterations in brain regions posited to subserve adolescent risk-taking, including the prefrontal cortex,^13–15,44^ insula,^15,45^ and striatum.^13,14^ Yet, previous work has mostly examined single metal exposures despite the fact that metals commonly co-occur in the environment and can jointly influence neurodevelopment and behavior.^46–49^ Examining associations between metal mixtures and child and adolescent brain and behavior is therefore essential for understanding the real-world impact of metal exposure on the developing brain.

Previous studies have shown that associations between early-life metal exposures and child and adolescent brain and behavior depend on both the timing and intensity of exposure.^13,14,33,50^ However, traditional biomarkers of early-life exposure (e.g., maternal blood, cord blood) capture only brief exposure periods and thus cannot resolve the timing of fetal and postnatal exposures.^25^ Previous work pioneered by Horton et al. (2018) and Rechtman et al. (2026) have overcome these limitations by using deciduous tooth biomarkers to link temporally resolved, longitudinal early-life metal exposure data with brain function and behavior. Deciduous teeth form in incremental layers known as “growth lines” beginning in the second trimester of pregnancy and continuing into infancy and early childhood.^51^ The neonatal line, an accentuated growth line formed at birth, demarcates pre- vs postnatally formed sections of teeth, allowing precise mapping of developmental windows.^52^ As metals incorporate into dentine during ongoing tooth mineralization, teeth provide a chronological record of exposure.^25,53–55^ By leveraging temporally resolved deciduous teeth biomarkers, we can identify specific developmental windows during which metal mixture exposure exerts the greatest influence on later-life brain function and behavior, and, ultimately, inform prevention and treatment strategies aimed at reducing adverse environmentally associated brain and behavioral outcomes.

In combination with temporally resolved deciduous tooth biomarkers, magnetic resonance imaging (MRI) offers a unique opportunity to examine the neural mechanisms through which metal exposure during specific early-life developmental windows may shape neurodevelopmental outcomes in adolescence. Relatively few studies have used MRI to examine the neurocognitive mechanisms of early-life metal exposure in adolescents.^13,14,16^ Investigating these mechanisms would provide insight into how such exposures may influence brain and behavioral outcomes relevant to adolescence, including risk-taking. Task-based functional magnetic resonance imaging (tb-fMRI) enables non-invasive measurement of in vivo neural activity during cognitive processes such as risky decision-making. Further, computational modeling approaches from neuroeconomics and decision sciences reveal latent cognitive processes underlying risky choice behavior (e.g., risk perception), that are often difficult to measure directly from behavioral task performance.^56,57^ Incorporating tb-fMRI and computational modeling approaches in children’s environmental health research therefore enables direct examination of how early-life environmental exposures shape neurocognitive mechanisms underlying complex behaviors such as adolescent risk-taking.

In this study, we used a tb-fMRI paradigm consisting of a risky decision-making task and a computational model of decision-making under risk to investigate how prenatal and postnatal exposure to 9 trace metals (barium (Ba), copper (Cu), lithium (Li), magnesium (Mg), manganese (Mn), lead (Pb), tin (Sn), strontium (Sr), and zinc (Zn)) is associated with neurocognitive correlates of risky decision-making in early adolescence. Building on prior evidence linking early-life metal exposure to alterations in brain regions and behaviors related to cognitive control and reward processing,^13–15,38–40,43,45,58,59^ we hypothesized that adolescents with higher metal mixture exposure would exhibit behavioral and neural profiles characteristic of individuals who engage in greater risk-taking.^60–63^ Specifically, we expected that adolescents with higher early-life exposure would show greater risk-taking tendency, risk sensitivity (i.e., risk seeking), and reward sensitivity (i.e., reward seeking), as well as reduced neural activation in cognitive control regions during risky decision-making and increased neural activation in reward regions during reward outcome processing.

## Materials and Methods

### Participants

The Programming Research in Obesity, Growth, and Social Stressors (PROGRESS) study is a longitudinal birth cohort investigating associations between early-life environmental exposures and child health outcomes in Mexico City.

Enrollment details are described in detail elsewhere.^64,65^ Briefly, between 2007 and 2011, pregnant women receiving prenatal care through the Mexican Social Security Institute were approached for enrollment. Eligibility criteria included: being < 20 weeks pregnant, being >18 years old, planning to remain in Mexico City for 3 years, phone access, and having no major health conditions or daily alcohol use. A total of 948 participants completed at least one study visit after giving birth.

Between 2018-2023, a subset of PROGRESS participants (n = 215) participated in a multimodal MRI study including structural and functional MRI. 214/215 adolescents completed the multimodal MRI scan, which included an fMRI-adapted version of the Cake Gambling Task (CGT).^61,62^ Behavioral analyses included participants with adequate behavioral task performance (n = 212). fMRI analyses included participants with adequate behavioral task performance, complete scanner timing information, and less than 1 voxel of motion (n = 185). Complete covariate data (child age, child sex, socioeconomic status) were available for all PROGRESS MRI participants. 190 participants had complete dentine metals data; Of these, 189 had adequate behavioral task data, and 166 had adequate fMRI data. Thus, the present study included 189 adolescents for behaviorally estimated outcomes (45.5% female, ages 8-14 years) and 166 adolescents for fMRI outcomes (45.2% female, ages 8-14 years).

Written informed consent was obtained from participants’ mothers and verbal assent was obtained from all participants after full discussion of study procedures. All study procedures were approved by the Institutional Review Boards (IRB) of Icahn School of Medicine at Mount Sinai, Harvard T. H. Chan School of Public Health, and the National Institute of Public Health Mexico and the National Institute of Perinatology, Mexico.

### Dentine metal biomarkers

As described previously,^16,25,53,55,66,67^ deciduous teeth free of defects (e.g., caries, extensive tooth wear) were collected and analyzed for metals using laser ablation-inductively coupled plasma-mass spectrometry (LA-ICP-MS; Agilent Technologies 8800 ICP-MS coupled with ESI 193 nm laser ablation unit). Teeth were washed and sectioned on a vertical (labial-lingual/ buccal-lingual) plane and the neonatal line and daily growth lines were identified to assign temporal information to sampling points. We identified 63 sampling points per tooth, spanning the second trimester of pregnancy (∼21 weeks gestation) to 43 weeks post birth (∼10 months postnatal), with a sampling frequency of approximately every 7–10 days. Metal intensities were normalized to calcium (Ca) and calibrated to NISH610 to account for variations in mineral density between samples and instrumental drift during analysis.

Metals with reliably detectable signals following background subtraction and normalization to Ca (Ba, Cu, Li, Mg, Mn, Pb, Sn, Sr, and Zn) were included in our analyses. Values were reported as a ratio of metal to Ca.

### Cake gambling task (CGT)

The present study used an fMRI-adjusted version of the Cake Gambling Task (CGT).^61,62^ The CGT is a child-friendly gambling paradigm adapted from the Cambridge Gambling Task, well-defined assessment of risky decision-making.^68,69^ Previous studies have used the CGT to study developmental differences in risk-taking.^61,62,70^ In each trial, participants chose between high-risk and low-risk gambles each associated with a probabilistic monetary reward. All information relevant for decision-making was presented in each trial, minimizing working memory demands. High-risk and low-risk gambles were displayed visually using a “cake” consisting of six wedges. Each wedge had two possible flavors: chocolate (brown), or strawberry (pink), presented in a 4:2 ratio or 5:1 ratio (Figure 1). Potential rewards associated with each gamble were displayed as stacked coins within either a pink or brown square located beneath the cake stimulus (Figure 1). Potential rewards associated with the low-risk gamble ranged from $1-3 Mexican pesos; potential rewards associated with the high-risk gamble ranged from $3-8 Mexican pesos. Selection of the majority flavor resulted in a high probability (67% <u>or</u> 83% chance) of obtaining a monetary reward (i.e., low-risk choice); selection of the minority flavor resulted in a low probability (17% <u>or</u> 33% chance) of a monetary reward (i.e., high-risk choice). The expected values (EV, probability x reward magnitude) for high-risk and low-risk gambles were varied such that the EV was not systematically higher for either type of gamble. High-risk choices were considered riskier because they were associated with a smaller probability of obtaining a reward and a greater variance in potential outcomes.

**Figure 1.**
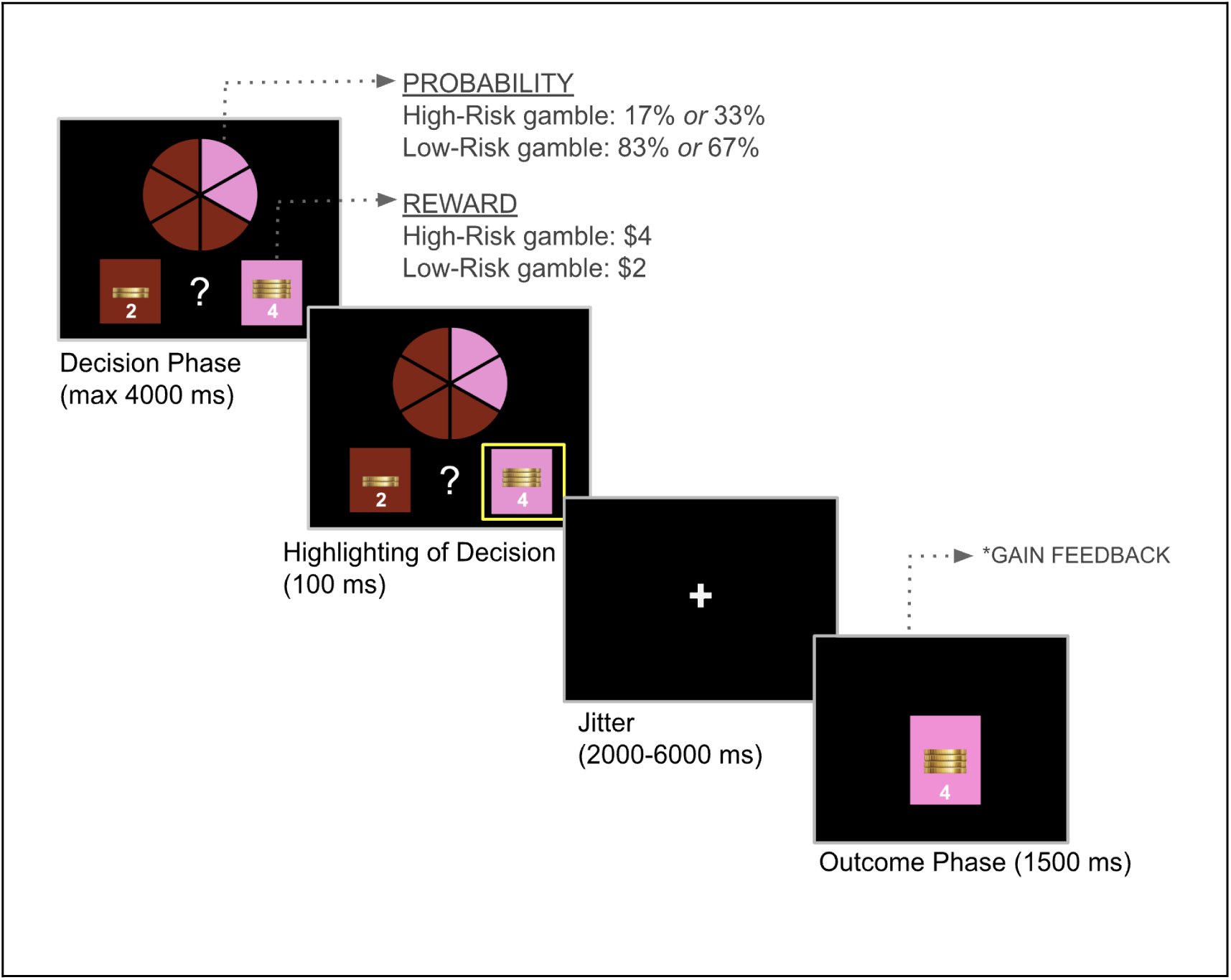
The Cake Gambling Task (CGT). Schematic depiction of a CGT trial. The high-risk decision (pink) was selected by the participant (highlighted box) and the associated reward ($4) was gained (gain feedback). *Adapted from Van Leijenhorst et al. (2010)*.

Each task trial had the following structure: First, the cake stimulus was presented, and participants had a maximum of 4000 milliseconds (ms) to select their choice by pressing their index or middle finger on a corresponding computer key. After participants selected their choice, the associated reward was highlighted for 100 ms and followed by a 2000-6000 ms fixation cross, during which a computer randomly selected one of the six wedges of cake. Participants were subsequently shown the outcome of the gamble for 1500 ms. The outcome screen displayed the result of the gamble (gain or no-gain) and the magnitude of the associated reward. For gain outcomes, participants were shown the coin stack associated with their choice. For no-gain outcomes, participants were shown this coin stack crossed out.

To encourage consistent engagement, participants were told that one of their choices during the task would be randomly selected to receive a prize, conveying that their choices and rewards on each trial are independent of each other, with no cumulative effect (e.g., coins are not accumulated across consecutive trials). Because all decisions/information from prior trials had no relevance to subsequent trials, the CGT has relatively low demands on executive functions that contribute to decision-making (e.g., working memory), making it less complex than other risky decision-making tasks (e.g., Iowa Gambling Task), and a robust measure of reward-based risky decision-making.^62,71^ High-risk decisions during the CGT have been positively associated with self-reported substance use^72^ and sensation-seeking behavior,^62^ suggesting that performance on the CGT may be associated with real-life risk-taking tendencies.

### Behavioral analysis: Risk-return decomposition

A risk-return framework^63,73,74^ was used to estimate the effect of changing returns (expected value) and changing risks (outcome variability) on the likelihood of selecting the high-risk vs the low-risk choice. Return (operationalized as the expected value (EV) of a decision’s outcome) and risk (operationalized as the standard deviation (SD) of the distribution of possible outcomes associated with a decision), were defined as follows:

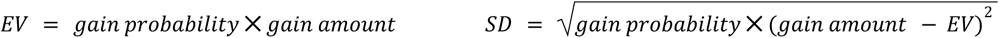

Risk (SD) and return (EV) for the high-risk choice were included as independent variables in the behavioral analysis, and the binary choice for the high-risk vs low-risk gamble served as the dependent variable. Correlations between risk and return for high-risk and low-risk gambles were modest, indicating minimal multicollinearity (high-risk gambles: r = 0.27; low-risk gambles: r = 0.40). We modeled participants’ choice behavior using a generalized linear mixed-effects model (GLMM) implemented in the lme4 package^75^ in R 4.2.2.^76^ Risk and return variables were centered and scaled and the unit of analysis was the binary decision level (high-risk choice/low-risk choice). The GLMM is given by:

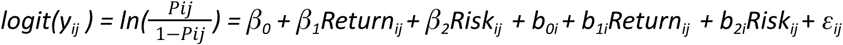

In which the log-odds of selecting the high-risk vs. low-risk gamble is estimated for each participant. *y_ij_* indicates the binary choice of the *i*th participant in the *jth* trial, with *yij = 1* denoting selecting the high-risk gamble *and yij = 0* denoting selecting the low-risk gamble. *Pij* denotes the probability of selecting the high-risk gamble (yij = 1) for the *i*th participant on the *j*th trial. Parameters in the model include: the fixed effects (*β_0_*, *β_1_*, and *β_2_*), participant-specific random effects *(b_0i_*, *b_1i_*, and *b_2i_*), and residual errors (*ε_ij_*). Given both risk and return were centered, the fixed intercept (*β_0_*) signified the tendency to select the high-risk gamble for the average risk and return.

Participant-specific random slopes for risk (*b_2i_*) and return (*b_1i_*) captured individual differences in sensitivity to risks and returns. Participant-specific random intercepts (*b_0i_*) captured individual differences in the tendency to select the high-risk gamble for average risk and return. Using this modeling approach,^63^ we decomposed risky choice-related processes into three distinct components: (1) risk-taking tendency (i.e., defined by the random intercept, or the overall tendency to select the high-risk gamble for average risk and return); (2) risk sensitivity (i.e., defined by random risk/SD slope), and 3) reward sensitivity (i.e., defined by random return/EV slope). Importantly, risk and reward sensitivity could be approach-related (i.e., positive coefficient denotes an increased likelihood of selecting the high-risk gamble with increasing risk/return), or avoidance-related (i.e., negative coefficient denotes a decreased likelihood of selecting the high-risk gamble with increasing risk/return). Given our hypothesis that early-life metal exposure increases susceptibility to maladaptive (i.e., excessive) adolescent risk-taking, we focused on approach-related risk sensitivity and reward sensitivity (i.e., risk seeking, reward seeking).

### MRI and fMRI data acquisition

MRI scans were acquired using a Philips Achieva 3T scanner equipped with an 8-channel head coil (Sense Head 8) at the Centro Nacional de Investigación en Imagenología e Instrumentación Médica (Ci3M), located within the Metropolitan Autonomous University (Universidad Autónoma Metropolitana, UAM) in Mexico City. All participants were shown an IRB-approved video about the MRI procedure prior to the scan and were accompanied by a trained adolescent psychologist throughout the entire scan. Participants received training on the CGT prior to the MRI scan. Anatomical T1-weighted scans were acquired for registration purposes using an MPRage sequence (301 volumes, TR = 7.5 ms, TE = 3.5 ms, FOV = 25×25 cm, Matrix =228 x 228, slice thickness = 1.2 mm). Participants watched an age-appropriate cartoon video while T1 scans were collected. Functional data were acquired using a field-echo-EPI gradient pulse (TR= 2000 ms, TE = 27 ms, slice thickness=3 mm, 37 axial slices, ascending slice acquisition, FOV=22 cm, Matrix size=80×80). A total of 48 trials were presented over the course of two event-related scans.

### fMRI data analysis

#### Image preprocessing

Image preprocessing was performed using Statistical Parametric Mapping 12 (SPM12) (http://www.fil.ion.ucl.ac.uk/spm/software/spm12/) and MATLAB R2022b (MathWorks). Preprocessing included slice timing correction, motion correction (realignment), co-registration, segmentation of structural images, spatial normalization using the anatomical image and the Montreal Neurological Institute (MNI) 305 template,^77^ and smoothing using a Gaussian kernel of 5 mm full-width at half maximum (FWHM).

#### fMRI statistical analyses

Statistical analyses were performed on participants’ data using the general linear model (GLM) implemented in SPM12. For each participant, the fMRI time series were modeled as a series of events convolved with a canonical hemodynamic response function (HRF). The decision and outcome phases of each trial were modeled as separate events in a single GLM. The GLM included an intercept, 6 motion parameters, and discrete regressors for decision events (high-risk choices, low-risk choices) and outcome events (gain outcomes, no-gain outcomes). Decision regressors were modeled as boxcar functions time-locked to the onset at which participants could make a decision (i.e., onset of the cake stimulus) and their duration was modeled by the choice reaction time for the respective trial. Outcome regressors were modeled as zero-duration stick functions time-locked to onset of the feedback presentation (i.e., gain or no-gain feedback). Decision analyses compared high-risk and low-risk gambles and outcome analyses compared gain and no-gain outcomes. The least-squares parameter estimates of the height of the best-fitting canonical HRF for each condition were used in pairwise contrasts. The resulting contrast images, computed on a participant-by-participant basis, were submitted to group analyses. At the group level, whole-brain contrasts between conditions were computed by performing one-sample t-tests. The following contrasts were computed: High-Risk > Low-Risk decisions, and Gain > No-Gain outcomes. All whole-brain analyses were corrected for multiple comparisons using family wise error (FWE) correction (p < .05 at the cluster level, with a cluster forming threshold of p < .001).

#### Region of interest (ROI) analysis

We used the MarsBaR toolbox implemented in SPM12^78^ to elucidate patterns of activation in significant clusters identified in the whole-brain analyses, focusing on regions previously linked to both adolescent risk-taking and early-life metal exposure.^13–15,45,58,59,79–82^ We selected 5 functional regions of interest (ROIs) from the following contrasts: 1) High-Risk > Low-Risk decisions: left and right angular gyrus/inferior parietal lobule; 2) Gain > No-Gain outcomes: left insula/rolandic operculum, left putamen/pallidum, bilateral precentral gyrus/postcentral gyrus/supplementary motor area. ROIs were defined as clusters that passed the p < .05, FWE-corrected threshold. Anatomical masks were created for each functional ROI using MarsBar. For functional ROIs that spanned several anatomical regions, we extracted the overlapping functional activation with anatomical regions defined by the Automated Anatomical Labeling (AAL)^83^ and Brodmann atlases implemented in MRIcron (https://www.nitrc.org/projects/mricron). Mean contrast estimate values were extracted from each functional ROI and modeled as outcomes in lagged weighted quantile sum (LWQS) regression analyses.^84^ MRIcroGL software (https://www.nitrc.org/projects/mricrogl) was used to visualize whole-brain activation clusters and functional ROIs.

### Statistical analyses

#### Covariates

Based on literature on metal exposure and adolescent risk-taking,^61,63,85–96^ we included the following covariates: child sex, child age at MRI, and socioeconomic status (SES). SES (lower, medium or higher) was calculated based on an index created by the Mexican Association of Market and Public Opinion Research Agencies (Spanish acronym AMAI).^97^ The AMAI index used 13 variables derived from a questionnaire (education of the head of household, number of rooms, number of bathrooms with showers, type of floor, number of light bulbs, ownership of car, hot water, automatic washing machine, videocassette recorder, toaster, vacuum cleaner, microwave oven, and personal computer) to classify individuals into six socioeconomic levels, which we further simplified into a relative three-level index of lower, medium and higher. This multidimensional index was preferred over single-indicator SES proxies (e.g., maternal education) given its broader capture of material deprivation and home environment factors relevant to neurodevelopmental outcomes,^98,99^ and its ability to more comprehensively account for the shared variance among correlated SES indicators that may confound exposure-outcome associations.

#### Descriptive statistics

Visual inspection was used to characterize dentine metal concentrations. Weekly dentine metal concentrations (19 weeks pre-birth to 43 postnatal weeks) were visualized using scatterplots implemented in the ggplot2 package^100^ in R.4.2.2.^76^ Chi-Square or Kruskall-Wallis tests were used to assess differences in the distribution of covariates (child sex, SES, maternal education at enrollment, gestational age at birth) between participants included in the present study, the PROGRESS MRI study, and the PROGRESS parent study (age 10-15 follow-up) (Supplementary Material - Table S1). Descriptive statistics were performed using R 4.2.2.^76^

#### Lagged weighted quantile sum (LWQS) regression

To examine time-varying associations between early-life metal mixture exposure and neurocognitive correlates of adolescent risky decision-making, we applied lagged weighted quantile sum (LWQS) regression models.^84,101^ Methods and procedures have been described in detail by Rechtman et al. 2026. Briefly, the LWQS integrates time-varying generalized weighted quantile sum regression (WQS)^102^ indices within the reverse distributed lag model (rDLM) approach^103^ to estimate time-varying associations between high-dimensional correlated mixtures and an outcome of interest. Discrete WQS models are estimated within adjacent time intervals (e.g., weeks) to construct a time-varying weighted index. The association between longitudinal WQS indices and an outcome of interest is elucidated through a smoothed association plot while adjusting for intra-participant observations.

Critical windows are identified as periods where the 95% confidence intervals for the time-varying association parameter (β_1_(t)) do not cross zero. In the case of a continuous outcome (e.g., neural activation), when the estimate of β_1_(t) is positive and significant, the mixture is positively associated with the outcome, and vice versa. Because some time intervals had a lower sample size relative to the number of mixture components, we implemented the random subset WQS^104^ in all LWQS models, in which subsets of components in the mixture were randomly selected multiple times (e.g. 1000 iterations) and averaged to estimate the weighted contribution of each component in the mixture. Models were estimated across 1000 random subsets, with 3 metals per subset. Metal concentrations were ranked in quartiles to estimate the weights of components contributing to the time-varying mixture index. The weighted association of each individual metal with the outcome was calculated by multiplying the weights at each time point (i.e., week) by the corresponding β_1_(t).Thus, unlike traditional WQS weights which are constrained to sum to one, weighted associations sum to the overall β1(t) at each timepoint and reflect each metal’s contribution to both the magnitude and direction of the overall mixture effect. For all analyses, the directionality of the association of the WQS index was constrained in the direction of the hypothesized harmful effects of the metal mixture in relation to adolescent risk-taking. Thus, associations were constrained in the positive direction for behaviorally estimated measures (risk-taking tendency, risk sensitivity, reward sensitivity) and reward-related brain activation, and in the negative direction for risk-related brain activation. All models were adjusted for covariates (child age, child sex and SES) and were performed using the lwqs and gamm4 packages^105,106^ in R 4.2.2.^76^

### Sensitivity analyses

#### Individual Metal Associations

Single Metal Analyses. In addition to the LWQS mixture analyses, we used rDLMs to examine associations between individual metal concentrations and select cognitive and neural outcomes, focusing on outcomes with the highest effect sizes from the mixture analyses. Analyses were performed using the gamm4 package^106^ in R 4.2.2.^76^

#### Construct Validity of Neurocognitive Outcomes

Brain Behavior Correlations. We used Spearman’s rank correlations to examine relationships between participants’ behavioral and neural responses to risk and reward. Specifically, we examined correlations between: 1) risk sensitivity and risk-related brain activation (High-Risk > Low-Risk decisions contrast) and 2) reward sensitivity and reward-related brain activation (Gain > No-Gain outcomes contrast). Correlations were plotted using the “stat_cor” function from the ggpubr package^107^ in R 4.2.2.^76^

Influence of Choice Variability on Neural Responses during High-Risk vs Low-Risk Decisions. To assess potential confounding by choice variability,^108–110^ we computed a group-level contrast for high-risk vs low-risk decisions, including only participants who chose the high-risk gamble for ≥ 20% of trials. The whole-brain analysis was similarly corrected for multiple comparisons using FWE correction (p < .05 at the cluster level, with a cluster forming threshold of p < .001).

## Results

### Demographic characteristics

Table 1 presents demographic characteristics of participants included in the the PROGRESS MRI study (n = 215), stratified by behavioral and fMRI subsets. Comparative demographic characteristics for the present subsets, the PROGRESS-MRI cohort, and the larger PROGRESS follow-up sample are reported in the Supplementary Material (Table S1). Participants’ mean age at MRI was 12.7 years (SD = 1.49) and 46.5% of participants were female.

Demographic characteristics (SES, child sex, maternal education and age at enrollment, and gestational age at birth) did not differ between MRI participants and non-MRI participants (Table S1). A flowchart of participant inclusion and exclusion criteria is shown in Supplementary Figure S1.

### Exposure characteristics

To characterize early-life metal exposure, we constructed scatterplots depicting weekly variations in dentine metal concentrations across the perinatal (prenatal and infancy) period, following the approach described in Rechtman et al. (2026). Figure 2 depicts temporal maps of the dentine metal mixture (Ba, Cu, Li, Mg, Mn, Pb, Sn, Sr, and Zn) between 19 weeks pre-birth (∼21 weeks gestation) and 43 weeks postnatally in PROGRESS MRI participants with adequate behavioral task data (n = 189). The figure demonstrates the variability in individual metal concentrations over time. Consistent with previous findings,^13,16,33^ dentine Mn levels were highest in the second trimester and declined steeply over the prenatal period. Levels continued to decline at a slower rate after birth. Dentine Zn levels slightly declined from the late prenatal period (∼3rd trimester) to the first several weeks after birth, with levels stabilizing thereafter. Dentine Ba and Sr levels similarly demonstrated a slight increase after birth that continued throughout the postnatal period. Dentine levels of Cu, Li, Mg, Pb, and Sn were relatively stable over the approximately 60-week sampling period. Identical trends were observed in participants with complete/adequate fMRI data (Supplementary Material - Figure S2). Together, these results suggest that metal concentrations show dynamic fluctuations over the perinatal period, with certain metals (e.g., Mn) exhibiting distinct temporal patterns.

**Figure 2.**
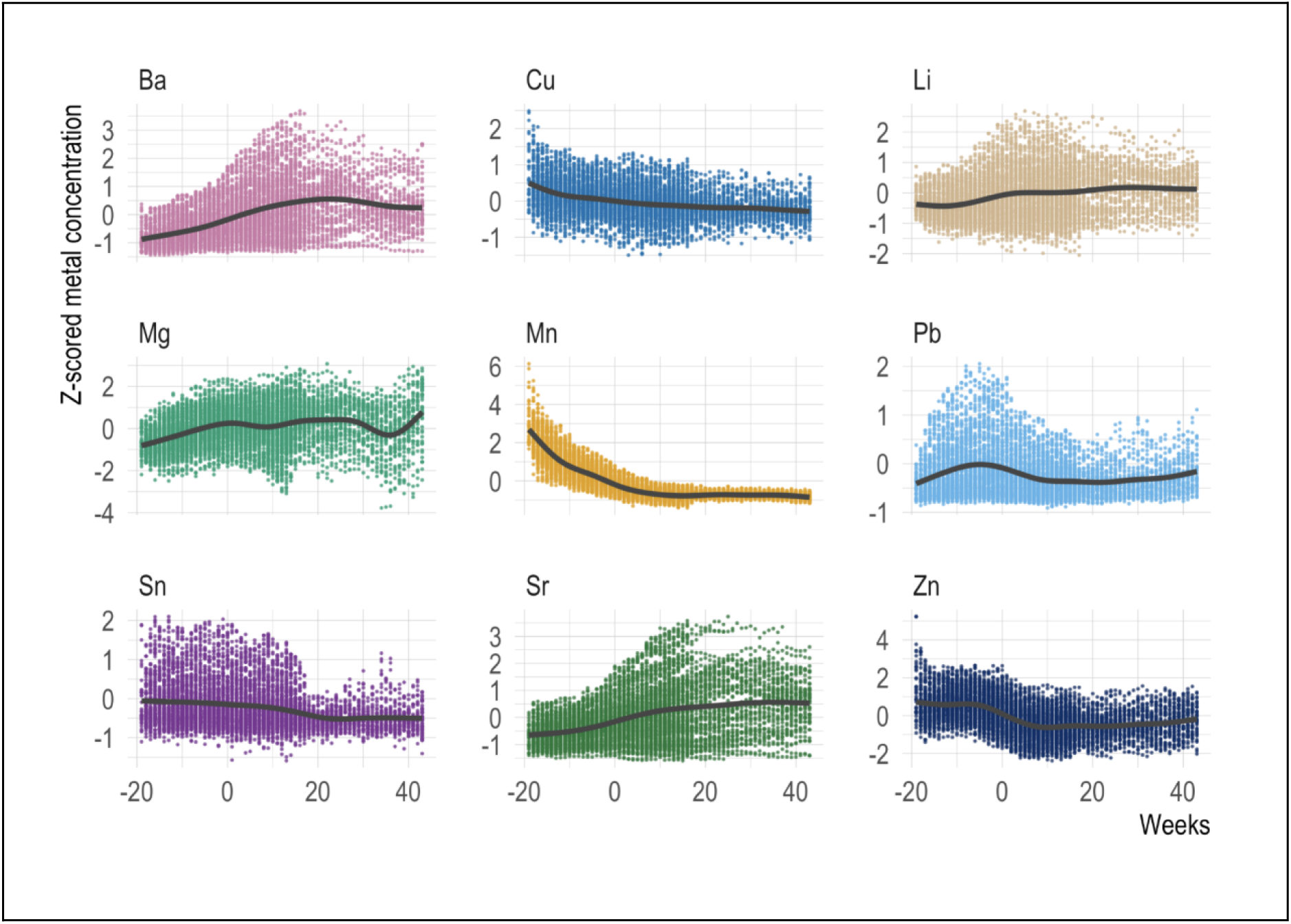
Individual dentine metal concentrations across development (from 19 weeks pre-birth to 43 weeks postnatally) in adolescents with adequate behavioral task data (n = 189). *<u>Note</u>*: Individual concentrations of dentine metals in PROGRESS MRI participants with complete dentine metals and adequate behavioral task data (n = 189). Colored dots represent individual tooth measurements for participants with approximately 60 measurements per participant. Lines represent locally estimated scatterplot smoothing (LOESS) curves. Outliers were excluded from the plot to improve visualization. The Y-axis shows z-scored metal concentrations normalized to Ca. The X-axis displays weekly concentrations from 19 weeks pre-birth through 43 weeks postnatally, with “0” representing birth. Metals include Barium: Ba, Copper: Cu, Lithium: Li, Magnesium: Mg, Manganese: Mn, Lead: Pb, Tin: Sn, Strontium: Sr, Zinc: Zn. *Adapted from Rechtman et al. (2026), with permission*.

### Cognitive constructs underlying adolescent risky decision-making

To examine the influence of changing risks and returns on adolescents’ risky choice behavior, we estimated group-level and participant-specific risk-taking tendency, risk sensitivity and reward sensitivity using a generalized linear mixed-effects model (GLMM) fit to participants’ decisions. Results from the GLMM revealed that at the group level (GLMM fixed effects), participants avoided risks and approached returns (Table S2); A higher risk was associated with a decreased likelihood of selecting the high-risk gamble (β = −0.133, p < 0.001), and a higher return was associated with an increased likelihood of selecting the high-risk gamble (β = 0.095, p < 0.001).

At the individual level (GLMM random effects), results revealed considerable variations in participants’ risk-taking tendency, risk sensitivity, and reward sensitivity, reflecting individual differences (Figure S3). Some participants exhibited greater risk-taking irrespective of changing risks and returns (i.e., a higher risk-taking tendency).

Additionally, some participants were more risk seeking (i.e., increasingly selected the high-risk gamble as risk increased), while others were more risk averse (i.e., decreasingly selected the high-risk gamble as risk increased). Participants similarly varied in reward sensitivity. Table 2 summarizes participant-specific estimates of risk-taking tendency, risk sensitivity, and reward sensitivity for the 189 participants included in the present study. Results from the table indicate that adolescents were overall risk-averse (risk-taking tendency, −0.80 ± 0.62; risk sensitivity, −0.13 ± 0.08) and reward-seeking (reward sensitivity, 0.09 ± 0.06). Together, these results suggest that adolescents show substantial individual differences in risk-taking, and in sensitivity to risk and rewards.

**Table 2.** GLMM (participant-specific) parameter estimates of risk-taking tendency, risk sensitivity and reward sensitivity among PROGRESS MRI participants included in the present study (n = 189)

| Cognitive Construct<br>(Parameter) | Mean ± SD |
| --- | --- |
| Risk-taking tendency<br>(random intercept) | $-0.80 \pm 0.62$ |
| Risk sensitivity<br>(random SD slope) | $-0.13 \pm 0.08$ |
| Reward sensitivity<br>(random EV slope) | $0.09 \pm 0.06$ |

*Note:* Participant-specific parameter estimates (random effects) from the generalized linear mixed effects model (GLMM) estimating the main effects of risk and return on adolescents’ likelihood (log-odds) of selecting the high-risk gamble. Risk-taking tendency (random intercept) reflects individual differences in the log-odds of selecting the high-risk gamble for average risk and return. Risk sensitivity (random SD slope) and reward sensitivity (random EV slope) reflects individual differences in how risk and return influence the log-odds of selecting the high-risk gamble.
EV, expected value

### Neural constructs underlying adolescent risky decision-making

#### Brain regions involved in high-risk vs low-risk decisions

To identify neural correlates of risky decision-making, we examined whole-brain activation patterns in response to high-risk vs low-risk decisions. The analysis revealed a cluster of activation in the left inferior parietal lobule (including the angular and supramarginal gyri), extending into the left occipital and temporal cortices (Table S3, Fig. S4). Two separate clusters of activation were observed in the left middle and superior temporal gyri, and the right inferior parietal lobule, extending into the right temporal cortex. The inferior parietal lobule has previously been linked to both early-life metal exposure^58^ and risky decision-making in adolescents.^79^ Thus, we selected the left and right angular gyrus/inferior parietal lobule (AG/IPL) as functional regions of interest (ROIs) to examine associations between risk-related brain activation and early-life metal mixture exposure (Figure 3).

**Figure 3.**
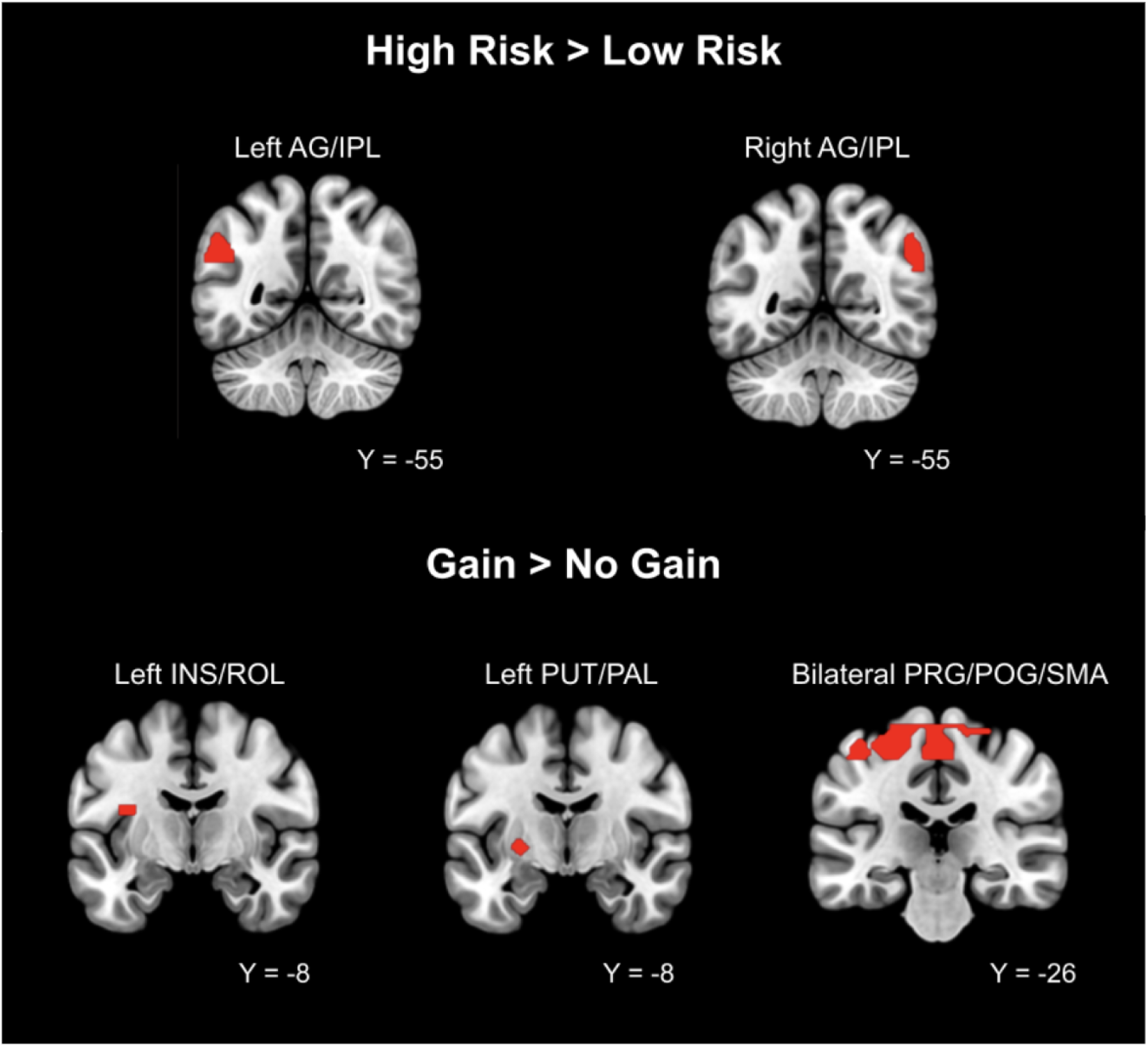
Functional regions of interest (ROIs) selected from whole-brain analyses for the High-Risk > Low-Risk decisions and Gain > No-Gain outcomes contrasts. ROIs were selected based on prior literature, focusing on regions linked to both adolescent risk-taking and early-life metal exposure. <u>Note</u>: AG: angular gyrus; IPL, inferior parietal lobule; INS, insula; ROL, rolandic operculum; PUT, putamen; PAL, pallidum; PRG, precentral gyrus; POG, postcentral gyrus; SMA, supplementary motor area

#### Brain regions involved in the processing of gain vs no-gain outcomes

To identify neural correlates of reward outcome processing, we examined whole-brain activation patterns in response to gain vs no-gain outcomes. The analysis revealed a cluster of activation in the bilateral precentral/postcentral gyri (including the paracentral lobule) that extended to additional regions within the parietal and frontal cortices (Table S3, Fig. S5). A cluster of activation was also observed in the left insula/rolandic operculum, which extended into the left basal ganglia (putamen, pallidum), and the parietal and temporal cortices. Additional clusters of activation included the left basal ganglia (putamen, pallidum), the right superior temporal gyrus, and the bilateral occipital cortex, with the latter activation extending into the left cerebellum. The analysis therefore revealed neural activation in brain regions previously linked to early-life metal exposure^13–15,45,59^ and reward processing in adolescents.^80–82^ We selected the following functional ROIs to examine associations between reward-related brain activation and early-life metal mixture exposure: left insula/rolandic operculum (INS/ROL), left putamen/pallidum (PUT/PAL), and bilateral precentral gyrus/postcentral gyrus/supplementary motor area (PRG/POG/SMA) (Figure 3).

### Effects of early-life metal mixture exposure on cognitive constructs underlying adolescent risky decision-making

To examine time-varying associations between early-life metal mixture exposure and cognitive constructs underlying adolescent risky decision-making, we modeled participant-specific risk-taking tendency, risk sensitivity and reward sensitivity as outcomes in LWQS regression models.

*Risk-taking tendency.* We observed a critical window of susceptibility spanning 12–43 weeks postnatally during which metal mixture exposure was significantly associated with increased risk-taking tendency (Figure 4A; maximum β = 0.18 [95% CI 0.040, 0.312]). At 43 weeks postnatally (i.e., the week with the strongest association in this window), a one-quartile increase in the mixture was associated with a 0.18 standard deviation increase in risk-taking tendency. This association was driven mainly by Pb, Sr, Zn and Li, in the early to mid-postnatal period (12–29 postnatal weeks) and by Li, Sr, Pb and Mn, in the late postnatal period (30–43 postnatal weeks) (Figure 4B).

**Figure 4.**
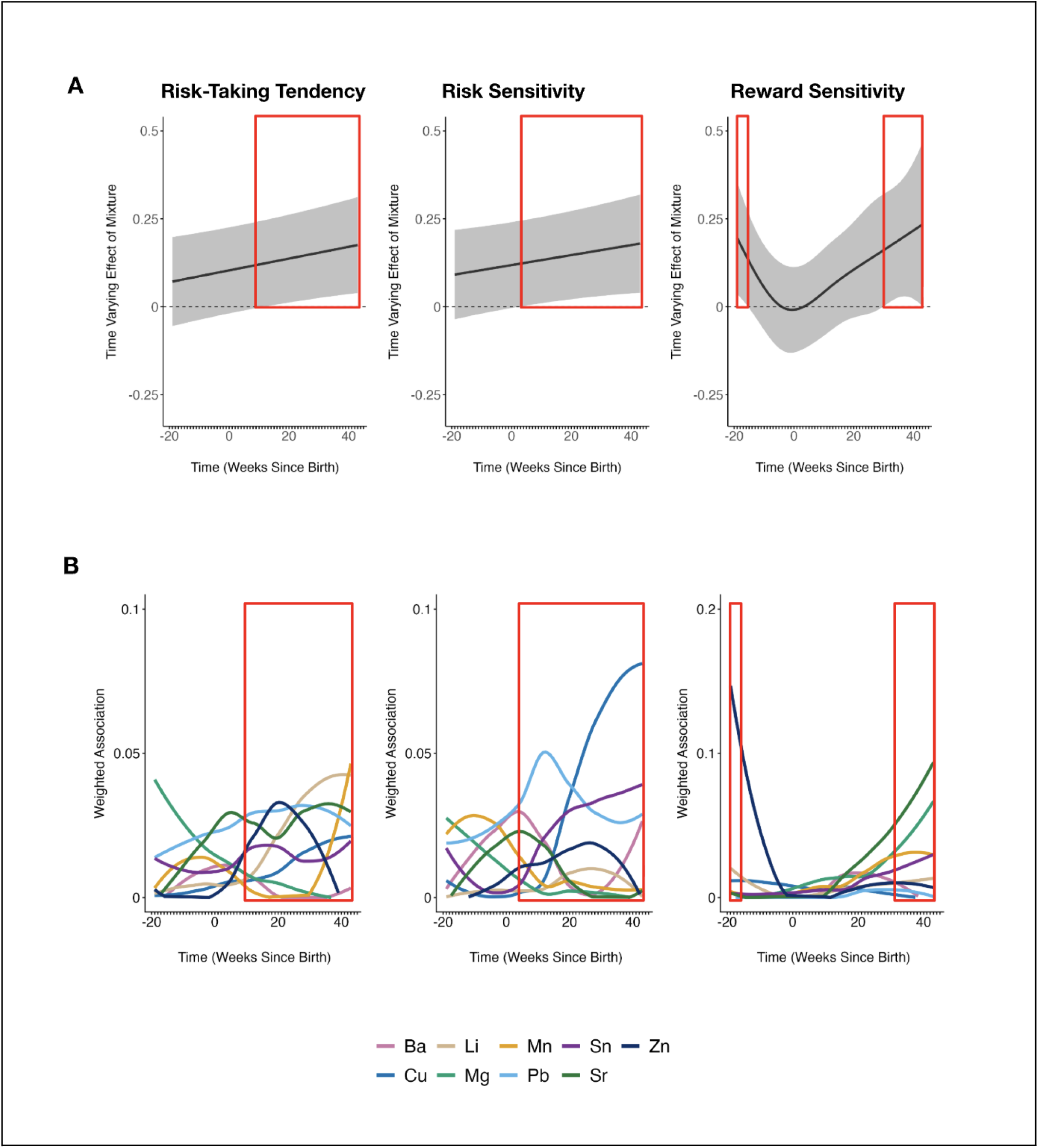
Time-varying associations between early-life metal mixture exposure and participant-specific risk-taking tendency, risk sensitivity and reward sensitivity. **(A)** Results from significant LWQS models for cognitive constructs derived from behaviorally estimated measures (N = 189). Gray shadows display 95% piecewise confidence intervals. The Y-axis represents time-varying correlations between the metal mixture and behaviorally estimated measures; participant-specific risk-taking tendency, risk sensitivity, and reward sensitivity. **(B)** Weighted associations (metal weight × β1(t)) of the individual metals driving the observed mixture effects are shown in panel A. In both figures (A,B), the X-axis represents the time since birth, indicating the timing (in weeks) of tooth sampling. <u>Note</u>: Barium: Ba; Copper: Cu; Lithium: Li; Magnesium: Mg; Manganese: Mn; Lead: Pb; Tin: Sn; Strontium: Sr; Zinc: Zn. *Adapted from Rechtman et al. (2026), with permission*.

*Risk sensitivity.* We observed a critical window at 3–43 weeks postnatally, during which metal exposure was significantly associated with increased risk sensitivity (Figure 4A; maximum β = 0.18 [95% CI 0.040, 0.319]). At 43 weeks postnatally (i.e., the week with the strongest association in this window), a one-quartile increase in the mixture was associated with a 0.18 standard deviation increase in risk sensitivity. This association was driven mainly by Pb, Ba, Sr and Sn in the early postnatal period (3–14 postnatal weeks); Pb, Cu, and Sn in the mid-postnatal period (15–29 weeks); and Cu in the late postnatal period (30–43 weeks) (Figure 4B).

*Reward sensitivity.* We observed two critical windows during which metal mixture exposure was significantly associated with increased reward sensitivity: 1) a mid-prenatal window spanning 19–16 weeks pre-birth (∼21–24 weeks gestation, maximum β = 0.20 [95% CI 0.037, 0.355]) and 2) a late postnatal window spanning 30–43 weeks postnatally (maximum β = 0.23 [95% CI 0.002, 0.467]) (Figure 4A). At 19 weeks pre-birth (i.e., the week with the strongest association in this first window), a one-unit increase in the mixture was associated with a 0.20 standard deviation increase in reward sensitivity. This association was driven by Zn (Figure 4B). At 43 weeks postnatally (i.e., the week with the strongest association in this second window), a one-quartile increase in the mixture was associated with a 0.23 standard deviation increase in reward sensitivity. This association was driven by Sr, Mg, Mn and Sn (Figure 4B). Altogether, these results suggest that the mid-prenatal and early to late postnatal periods are critical exposure windows for the effect of metal mixture exposure on cognitive constructs underlying adolescent risky decision-making.

### Effects of early-life metal mixture exposure on neural constructs underlying adolescent risky decision-making

To examine time-varying associations between early-life metal mixture exposure and neural constructs underlying adolescent risky decision-making, we modeled mean contrast estimate values from the functional ROIs for high-risk vs. low-risk decisions (left and right AG/IPL), and gain vs. no-gain outcomes contrasts (left INS/ROL, left PUT/PAL, bilateral PRG/POG/SMA), as outcomes in LWQS regression models.

*High-Risk > Low-Risk decisions.* We observed critical windows during which metal mixture exposure was significantly associated with decreased activation in the left and right AG/IPL during high-risk vs. low-risk decisions. At 19–10 weeks pre-birth (∼21–30 weeks gestation), metal mixture exposure was significantly associated with decreased activation in the left AG/IPL (Figure 5A; maximum β = −0.19 [95% CI −0.364, −0.006]). At 19 weeks pre-birth (i.e., the week with the strongest association in this window), a one-quartile increase in the mixture was associated with a 0.19 standard deviation decrease in left AG/IPL activation. This association was driven primarily by Pb, Mg and Zn (Figure 5B). At 20–43 weeks postnatally, metal mixture exposure was significantly associated with decreased activation in the right AG/IPL (Figure 5A; maximum β = −0.24 [95% CI −0.407, −0.065]). At 34 weeks postnatally (i.e., the week with the strongest association in this window), a one-quartile increase in the mixture was associated with a 0.24 standard deviation decrease in right AG/IPL activation. This association was primarily driven by Sr (Figure 5B). Overall, these findings suggest that mid to late prenatal and postnatal periods are critical exposure windows for the effect of metal mixture exposure on risk-related brain activation.

**Figure 5.**
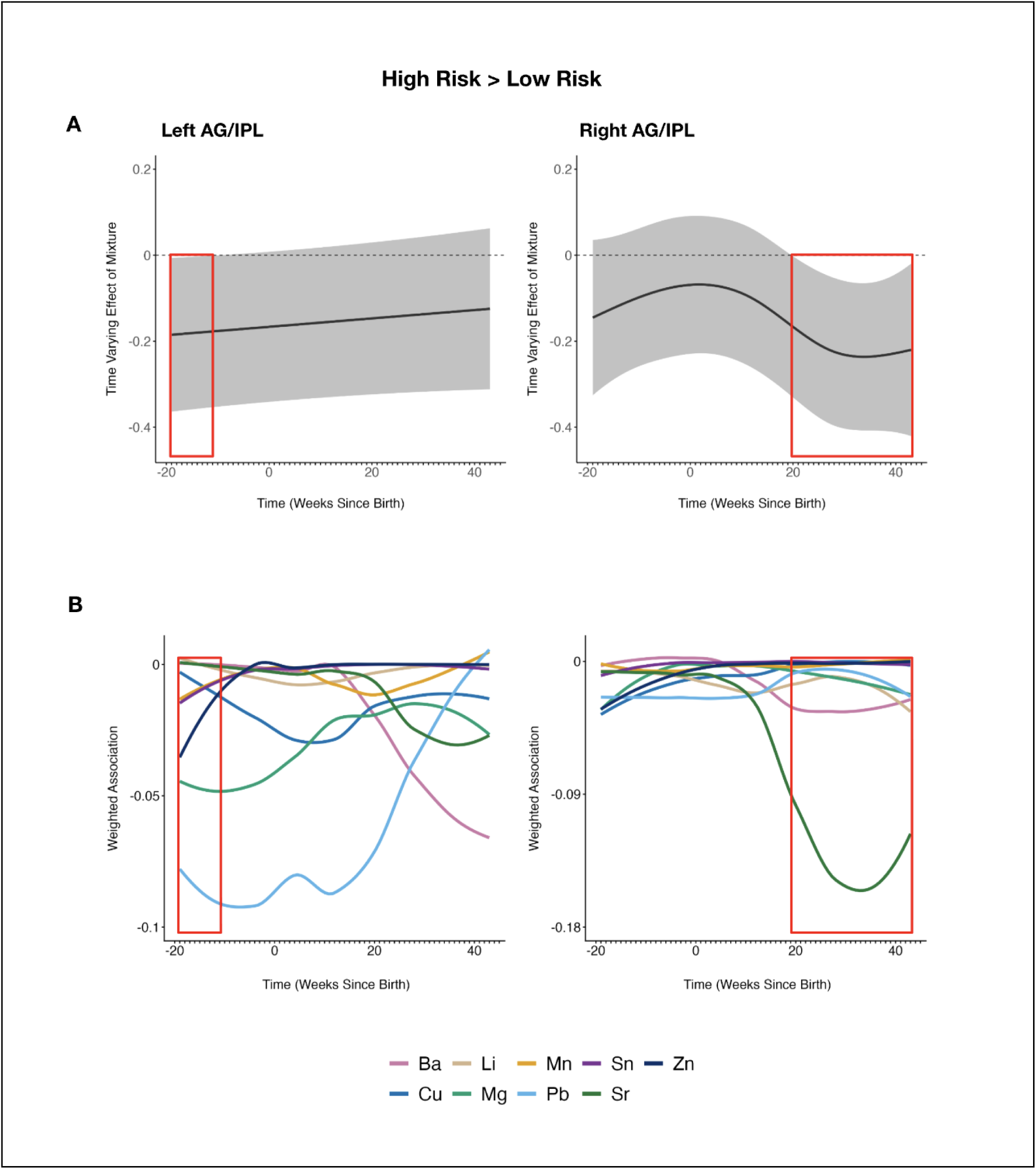
Time-varying associations between early-life metal mixture exposure and risk-related neural activation in functional regions of interest derived from the High Risk > Low Risk decisions contrast. **(A)** Results from significant LWQS models for neural constructs derived from the High-Risk > Low-Risk decisions contrast (N = 166). Gray shadows display 95% piecewise confidence intervals. The Y-axis represents time-varying correlations between the metal mixture and functional ROIs: left AG/IPL, right AG/IPL. **(B)** Weighted associations (metal weight × β1(t)) of the individual metals driving the observed mixture effects are shown in panel A. In both figures (A,B), the X-axis represents the time since birth, indicating the timing (in weeks) of tooth sampling. <u>Note</u>: AG/IPL: angular gyrus/inferior parietal lobule; Barium: Ba; Copper: Cu; Lithium: Li; Magnesium: Mg; Manganese: Mn; Lead: Pb; Tin: Sn; Strontium: Sr; Zinc: Zn

*Gain > No-Gain outcomes.* We observed critical windows in which metal exposure was significantly associated with increased activation in the left INS/ROL during gain vs no-gain outcomes. At 27–43 weeks postnatally, metal mixture exposure was significantly associated with increased activation in the left INS/ROL (Figure 6A; maximum β = 0.28 [95% CI 0.095, 0.470]). At 43 weeks postnatally (i.e., the week with the strongest association in this window), a one-decile increase in the mixture was associated with a 0.28 standard deviation increase in left INS/ROL activation. This association was driven by Mg (Figure 6B). No significant associations were observed between metal mixture exposure and reward-related activation in the left PUT/PAL and bilateral PRG/POG/SMA (Figure 7). Together, these results suggest that the late postnatal period is a critical exposure window for the effect of metal mixture exposure on reward-related brain activation.

**Figure 6.**
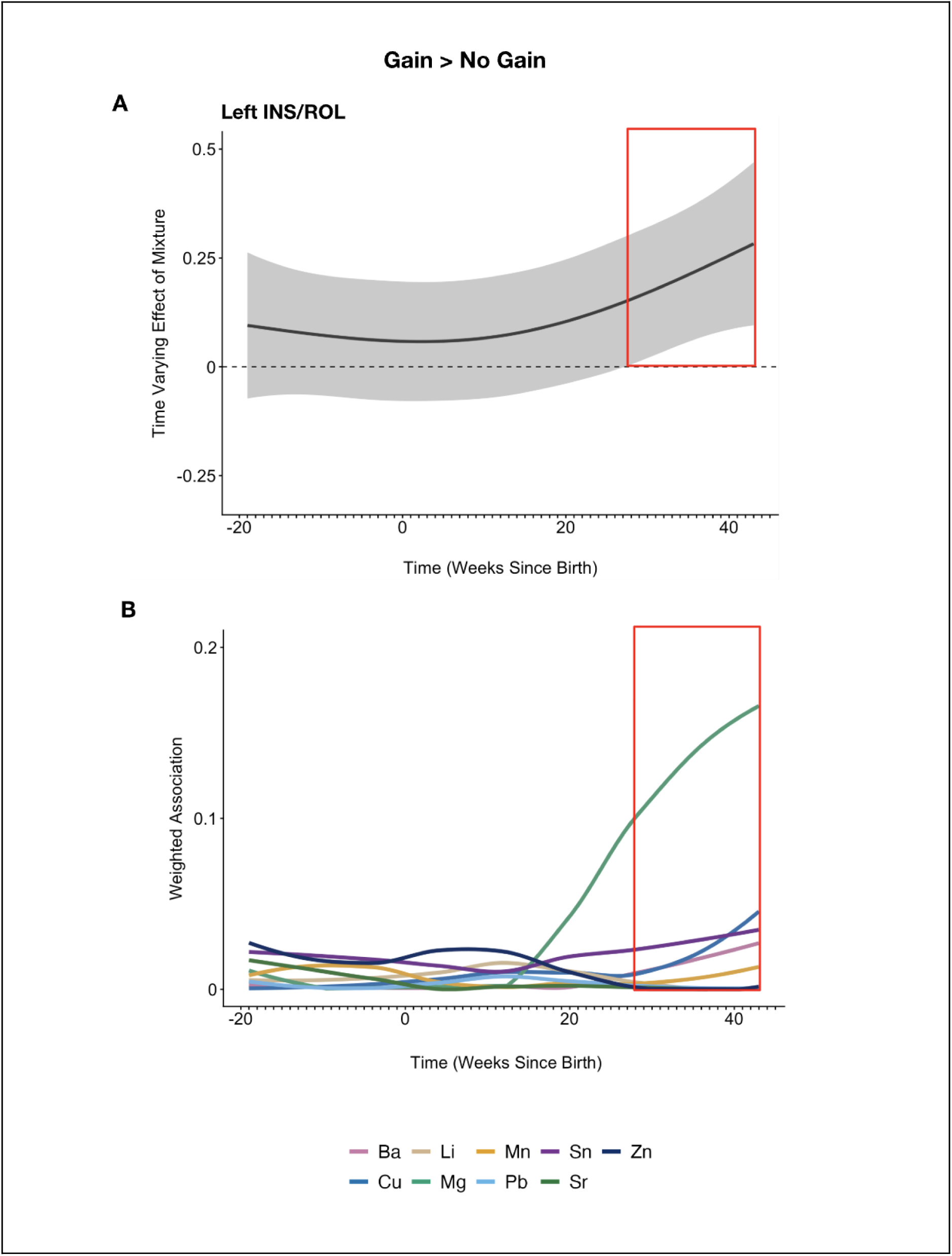
Time-varying associations between early-life metal mixture exposure and reward-related neural activation in functional regions of interest derived from the Gain > No-Gain outcomes contrast. **(A)** Results from significant LWQS models for neural constructs derived from the Gain > No-Gain outcomes contrast (N = 166). Gray shadows display 95% piecewise confidence intervals. The Y-axis represents time-varying correlations between the metal mixture and functional ROIs: left INS/ROL. **(B)** Weighted associations (metal weight × β1(t)) of the individual metals driving the observed mixture effects are shown in panel A. In both figures (A,B), the X-axis represents the time since birth, indicating the timing (in weeks) of tooth sampling. <u>Note</u>: INS/ROL: insula/rolandic operculum, Barium: Ba; Copper: Cu; Lithium: Li; Magnesium: Mg; Manganese: Mn; Lead: Pb; Tin: Sn; Strontium: Sr; Zinc: Zn

**Figure 7.**
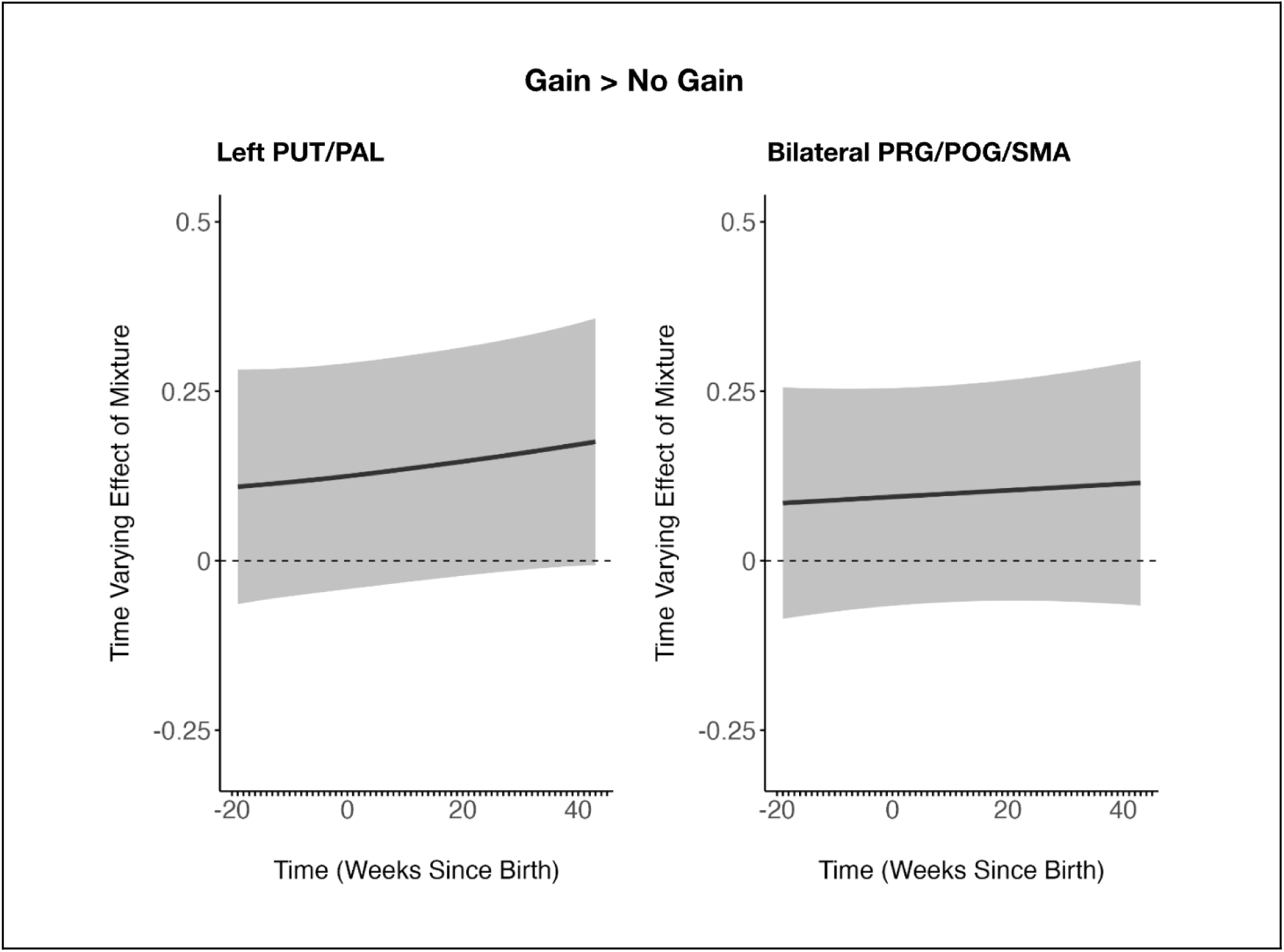
Non-significant time-varying associations between early-life metal mixture exposure and reward-related neural activation in functional regions of interest derived from the Gain > No-Gain outcomes contrast. Results from non-significant LWQS models for neural constructs derived from the Gain > No-Gain outcomes contrast (N = 166). Gray shadows display 95% piecewise confidence intervals. The Y-axis represents time-varying correlations between the metal mixture and functional ROIs: left PUT/PAL; bilateral PRG/POG/SMA. The X-axis represents the time since birth, indicating the timing (in weeks) of tooth sampling. Note: PUT/PAL: putamen/pallidum; PRG/POG/SMA: precentral gyrus/postcentral gyrus/supplementary motor area

### Sensitivity analyses

#### Individual Metal Associations

Single Metal Analyses. We used rDLMs to examine time-varying associations between individual metal concentrations and cognitive and neural constructs underlying adolescent risky decision-making. For cognitive constructs, we focused on reward sensitivity, as this outcome exhibited the highest maximum effect size. Results revealed a significant association between Mg exposure and reward sensitivity at 25–43 weeks postnatally (Fig. S6: maximum β = 0.39 [95% CI 0.106, 0.671]). No significant associations were observed between other metals and reward sensitivity. These results support the findings in the mixture analysis, suggesting that Mg contributes to the association between metal mixture exposure at 30–43 postnatal weeks and increased reward sensitivity. For cognitive control brain regions, we focused on the right AG/IPL, given this outcome had the highest maximum effect size. Results from the rDLMs were not significant (Fig. S7). Single metal analyses for reward brain regions focused on associations with the left INS/ROL (i.e., the region with the highest maximum effect size); the results from the rDLMs also revealed no significant associations (Fig. S8). Together, these findings suggest that the associations identified in the mixture analyses for neural activation in reward and cognitive control regions were likely driven by multiple metals within the mixture.

#### Construct Validity of Neurocognitive Outcomes

Brain Behavior Correlations. We examined relationships between participants’ behavioral and neural responses to risk and reward using Spearman’s rank correlations. We observed a significant negative correlation between risk sensitivity and left AG/IPL activation (Fig. S9; *ρ* = −0.28, p < 0.001). A similar, albeit non-significant correlation, was observed in the right AG/IPL (*ρ* = −0.13, p = 0.07). In contrast, no significant correlations were observed between reward sensitivity and reward-related brain activation (all *p*s > 0.4). Collectively, these findings align with previous literature suggesting a role of the posterior parietal cortex (including in the IPL) in decision-making under risk^.61,79,111–114^

Influence of Choice Variability on Neural Responses during High-Risk vs Low-Risk Decisions. To assess potential confounding by choice variability,^108–110^ we re-examined group-level activation for the High-Risk > Low-Risk decision contrast, including only participants who chose the high-risk gamble for ≥ 20% of the trials. Results revealed similar clusters of activation in the left and right AG/IPL (Fig. S10), suggesting that the findings for these ROIs in the initial analysis were not confounded by participants who showed low choice variability.

## Discussion

In the current study, using weekly-resolved metal profiles from children’s deciduous teeth, we identified pre- and postnatal exposure windows associated with individual differences in risk-taking tendency, risk sensitivity, and reward sensitivity. We also identified developmental windows during which metal mixture exposure was linked to reduced risk-related activation in cognitive control brain regions (bilateral angular gyrus/inferior parietal lobule) and greater reward-related activation in reward brain regions (left insula/rolandic operculum). Taken together, our findings suggest that the perinatal period spanning from the second trimester to 10 postnatal months is a sensitive developmental window during which mixed metal exposure influences neurocognitive correlates of adolescent risky decision-making. Additionally, associations between early-life metal mixture exposure and these neurocognitive correlates vary across pre- and postnatal development, highlighting the importance of considering exposure timing when estimating associations with later-life brain and behavioral outcomes.

Extending the teeth-based, time-varying analysis framework pioneered by Horton et al. (2018) and Rechtman et al. (2026), this is the first study to investigate time-varying associations between early-life metal mixture exposure and neurocognitive mechanisms underlying adolescent risky decision-making. Prior studies examining the role of environmental factors on adolescent risk-taking have primarily focused on concurrent social factors^115,116^ (e.g., parental dynamics, peer influence), leaving early-life susceptibility to environmental toxicant exposures understudied. Using a DOHaD framework and temporally resolved deciduous tooth biomarkers, we identified critical exposure windows overlapping with the timing of key neuromaturational processes throughout the second trimester to early infancy^22,23,117^ (e.g., neuronal proliferation, synaptogenesis, myelination). Disruption of these processes via metal exposure may negatively impact neurodevelopmental trajectories of cognitive control and reward processing, potentially increasing susceptibility to maladaptive adolescent risk-taking. Aligning with prior findings,^16,33^ results consistently implicated the postnatal window of 7–10 months as a critical window of exposure. This may be attributable to differential exposures in infancy, including direct exposures through dietary sources^118–125^ (e.g., breastmilk, infant formula, commercial baby food) and/or interactions with the environment^126,127^ (e.g., inhalation, dermal contact), which may lead to higher exposure levels compared to gestation. Further, the loss of protective mechanisms provided by the placenta,^128^ coupled with changes in metal metabolism,^129^ may also contribute to heightened vulnerability to metals in infancy. Blood levels of several metals have been found to be elevated in infants compared to neonates,^130^ supporting this notion. Altogether, our findings highlight the utility of temporally resolved exposure reconstruction via deciduous teeth, as these exposure windows may have been missed using traditional biomarkers typically measured at single pre- or postnatal time points. Further, teeth provide a direct measure of fetal exposure unlike proxies such as maternal blood and therefore minimize the risk of exposure misclassification.^25^

Our findings highlight the importance of examining metal mixtures rather than individual metals. Although prior studies have linked early-life exposure to individual metals, particularly Pb and Mn, to brain regions and behaviors implicated in adolescent risk-taking,^13–15,33–36,38–43^ far fewer have evaluated mixture effects. Epidemiologic and toxicologic evidence indicate that metals can interact synergistically or antagonistically through shared biological pathways and neurotoxic mechanisms (e.g., oxidative stress, neurotransmitter disruption).^131–133^ Previous studies have shown metal co-exposure to exacerbate neurodevelopmental and behavioral deficits in children and adolescents, with Pb and Mn most notably implicated.^46,47,49,134–136^ Further, experimental animal work has shown that Mn can alter the accumulation and retention of several metals within the brain (i.e., Pb, Cu, Zn)^131,137–140^ and that Cu can increase Pb accumulation.^141^ The mixture in the present study included essential (Mn, Zn, Cu, Mg) and non-essential (Li, Ba, Sr, Pb, Sn) metals, both of which were consistently found to be among the top contributors in the observed associations. Prior studies have linked joint exposures to essential and non-essential metals to alterations in child and adolescent neurobehavior,^46,142–144^ consistent with our findings. Such interactions likely contribute to the “mixture effects” we observed in the present study. Results from the single metal analyses support this; the majority of the critical exposure windows identified in the mixture analyses were not observed for individual metal associations, suggesting that associations within these windows are likely driven by the joint effect of multiple metals in the mixture. The only exception was Mg, which was positively associated with increased reward sensitivity at 30-43 postnatal weeks and emerged as a top contributor to the mixture effect. Mg has been shown to facilitate dopaminergic neurotransmission,^145,146^ a system heavily involved in reward processing,^147^ which may explain this association. Additionally, while substantial evidence indicates a protective role of Mg in relation to neurodevelopment,^148,149^ higher perinatal Mg exposure has also been linked to increased neuronal death in a neonatal rodent model,^150^ as well as an elevated risk of attention deficit/hyperactivity disorder (ADHD) and autism spectrum disorder (ASD) in children,^151^ suggesting that excess early-life Mg exposure—in addition to Mg deficiency—may also adversely impact neurodevelopment. Overall, our results suggest that examining metal mixtures provides a more accurate picture of the impact of early-life exposure on child and adolescent brain and behavior.

This study, to our knowledge, is also the first to integrate tb-fMRI and computational behavioral modeling with a measure of early-life metal mixture exposure. As such, these findings provide further insight into neurocognitive mechanisms that may underlie previously reported associations between early-life metal exposure and alterations in brain regions and behaviors implicated in adolescent risk-taking.^13–15,39,40,43,45,58,59^ Using a well-defined, age appropriate fMRI task of reward-based risky decision-making and a risk-return framework, we linked early-life metal mixture exposure to: a) individual differences in latent constructs underlying risky choice behavior and b) neural activity during risky decision-making and reward outcome processing. Consistent with previous neuroimaging studies of adolescent risky decision-making, we observed activation in regions associated with cognitive control (angular and supramarginal gyrus, inferior parietal lobule) and reward processing (basal ganglia, insula, primary and supplementary motor cortices).^61,79,80,152–154^ Furthermore, we observed that activation in the bilateral angular gyrus/inferior parietal lobule during high-risk vs low-risk decisions was negatively correlated with behaviorally estimated risk sensitivity, aligning with previous findings linking posterior parietal activity to individual differences in risk-taking preference^61^ and self-reported risk-taking.^111^ Interestingly, we did not observe significant correlations between neural activation during reward outcome processing and behaviorally estimated reward sensitivity. However, prior studies have linked reward-related activation in the brain regions identified in our fMRI analyses—most notably the insula and putamen—with sensation-seeking (a psychological correlate of reward sensitivity) in adolescence.^153,154^ Therefore, the present findings linking early-life metal exposure to activation in these regions are likely relevant in the context of adolescent risk-taking. In sum, this study builds upon previous work by linking early-life metal exposure with neurocognitive processes directly related to adolescent risky decision-making. Additionally, results from the mixture analyses confirm previous epidemiological findings suggesting early-life metal exposure may promote adolescent risk-taking through its impact on neural and cognitive constructs integral to decision-making under risk (e.g., parietal activation, reward sensitivity, inhibitory control).^14,15,38–40,43^ By highlighting these specific mechanisms, this study provides insight into potential interventions (e.g., neurocognitive/neurofeedback training) targeting metal-associated brain regions that may help mitigate susceptibility to maladaptive adolescent risk-taking.^155–159^

This study has several limitations that we acknowledge. First, studying early adolescents may have limited the ability to assess peak risk-taking, which typically occurs in mid to late adolescence.^6^ Second, the deciduous tooth biomarker only captures metal exposure from the second trimester onward, and similar to other biomarker-based assessments, may introduce some degree of exposure measurement error. However, measurement error in biomarker-based exposure assessments is typically assumed to be classical-type, which generally results in attenuation of effect estimates toward the null rather than inducing spurious associations.^160,161^ Third, the LWQS method does not enable the estimation of non-additive effects (i.e., metal-metal interactions). Additionally, as with all mixture analyses, the potential for bias amplification due to residual confounding by unmeasured variables warrants consideration.^162^ We also acknowledge the trade-off between personal (e.g., teeth, blood) and proxy-based (e.g., ambient air, drinking water) exposure assessments, as personal exposure measures may be more susceptible to residual confounding by individual-level behaviors and factors (e.g., prenatal tobacco exposure, maternal diet, and breastfeeding).^163^ However, direct evidence linking these factors to adolescent risk-taking behavior and associated neural activation is limited, leaving their potential for confounding the observed associations unclear. Future studies should incorporate measures of maternal and infant individual-level factors to elucidate their potential role as confounders or effect modifiers of these associations. Finally, the cross-sectional nature of the MRI data and the modest sample size limited the ability to examine developmental trajectories and detect sex-specific associations, respectively. Our findings therefore require cautious interpretation, particularly for associations with smaller effect sizes. Given the dynamic changes in brain development throughout adolescence, future studies should include longitudinal follow-up with a larger cohort of participants to further elucidate the relationship between early-life metal mixture exposure and neurocognitive correlates of adolescent risky decision-making. Moreover, as previous studies have observed sex differences in associations between metal exposure and risk-taking-related brain regions and behaviors,^13,164–166^ future studies could build upon our findings by investigating sex-specific effects.

## Conclusion

Using weekly-resolved metal profiles from children’s deciduous teeth, we identified pre- and postnatal windows during which early-life metal mixture exposure was associated with alterations in neurocognitive correlates of adolescent risky decision-making. Across these findings, the postnatal period of 7–10 months emerged as a critical window of susceptibility to metal mixture exposure, suggesting that infancy is a crucial developmental period during which exposures shape neural systems supporting adolescent risk-taking. By integrating task-based functional neuroimaging and computational behavioral modeling with a teeth-based LWQS approach previously applied to brain and behavioral outcomes (Horton et al. 2018; Rechtman et al. 2026), this study advances the understanding of environmental influences on the developmental origins of adolescent risk-taking and provides additional insight into underlying mechanisms of metal neurotoxicity. Collectively, the present findings underscore the importance of exposure timing and mixture effects when evaluating environmental influences on child and adolescent brain and behavior and highlight potential targets for prevention strategies aimed at improving neurodevelopmental outcomes.

## Data Availability

All data produced in the present study are available upon reasonable request to the authors

