## Supplementary Material for "Early-life critical windows of susceptibility to metal mixture exposure and neurocognitive mechanisms underlying adolescent risky decision-making"

**Table S1. Demographic characteristics of the adolescents included in the PROGRESS parent study (n = 581), the PROGRESS MRI study (n = 215), and the present study (n = 189, n = 166).**

| Characteristic | PROGRESS Parent[a] (n = 581) | PROGRESS MRI (n = 215) | CGT Behavior (n = 189) | CGT fMRI (n = 166) | p[b] |
|---|---|---|---|---|---|
| **SES** (%) Lower/Medium/Higher | 53.2/36.5/10.3 | 53.0/36.7/10.2 | 49.7/38.6/11.6 | 48.8/38.6/12.7 | 0.95 |
| **Child sex** (% Female) | 47.7% | 46.5% | 45.5% | 45.2% | 0.88 |

*CGT,* cake gambling task; *SES*, socioeconomic status

[a] PROGRESS parent cohort includes all participants who completed the age 10-15 follow-up: PROGRESS MRI subset (n = 215), non-MRI subset (n = 366)

[b] P-values reflect differences between the non-MRI subset, the PROGRESS MRI subset, and the behavioral and fMRI subsets included in the present study. Differences in the distribution of sex and SES were tested using Chi-Square tests.

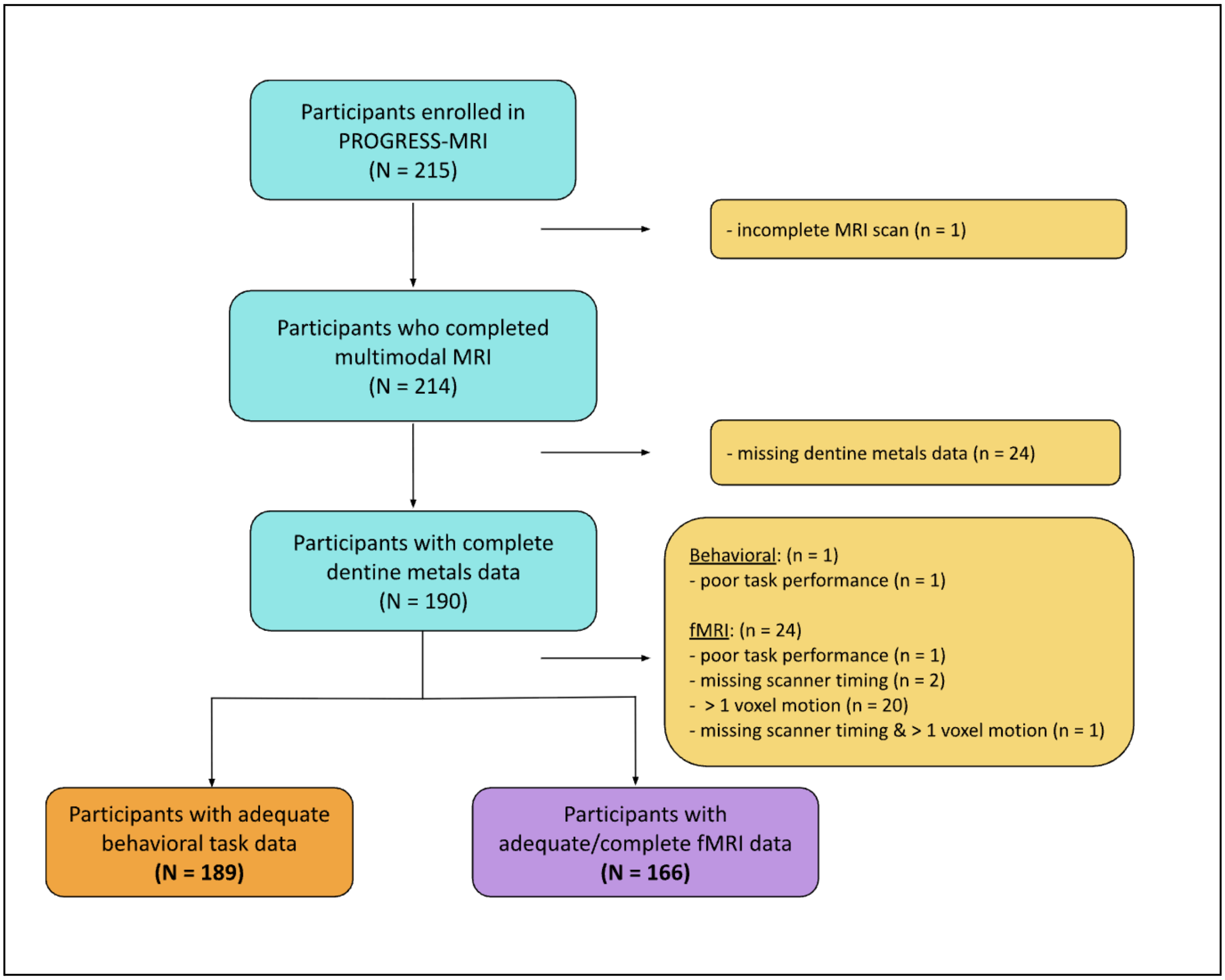


**Figure S1**. Flowchart of PROGRESS-MRI participant inclusion and exclusion criteria for the present study.

**Figure S2. Individual dentine metal concentrations across development (from 19 weeks pre-birth to 43 weeks postnatally) in adolescents with adequate/complete fMRI data (n = 166).**

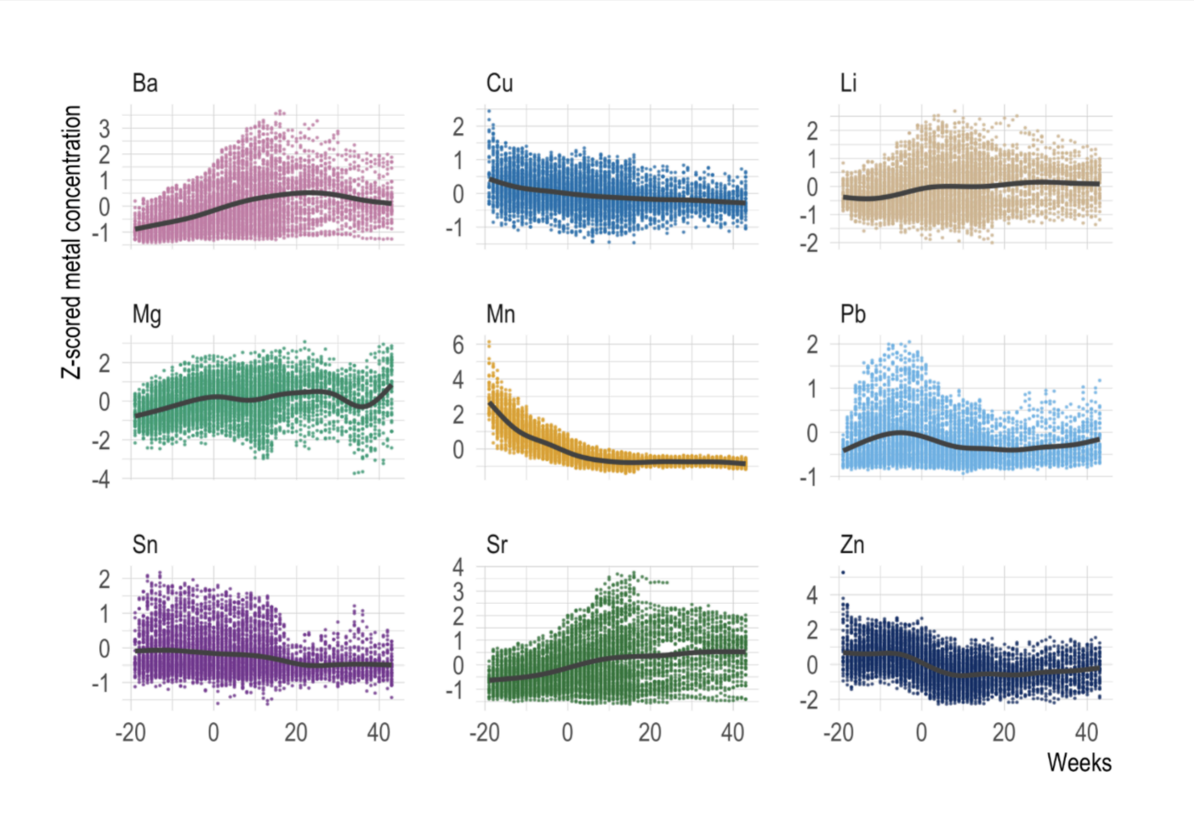


*Note*: Individual concentrations of dentine metals in PROGRESS MRI participants with complete dentine metals and fMRI data (n = 166). Colored dots represent individual tooth measurements for participants with approximately 60 measurements per participant. Lines represent locally estimated scatterplot smoothing (LOESS) curves. Outliers were excluded from the plot to improve visualization. The Y-axis shows z-scored metal concentrations normalized to Ca. The X-axis displays weekly concentrations from 19 weeks pre-birth through 43 weeks postnatally, with "0" representing birth. Metals include Barium: Ba, Copper: Cu, Lithium: Li, Magnesium: Mg, Manganese: Mn, Lead: Pb, Tin: Sn, Strontium: Sr, Zinc :Zn.

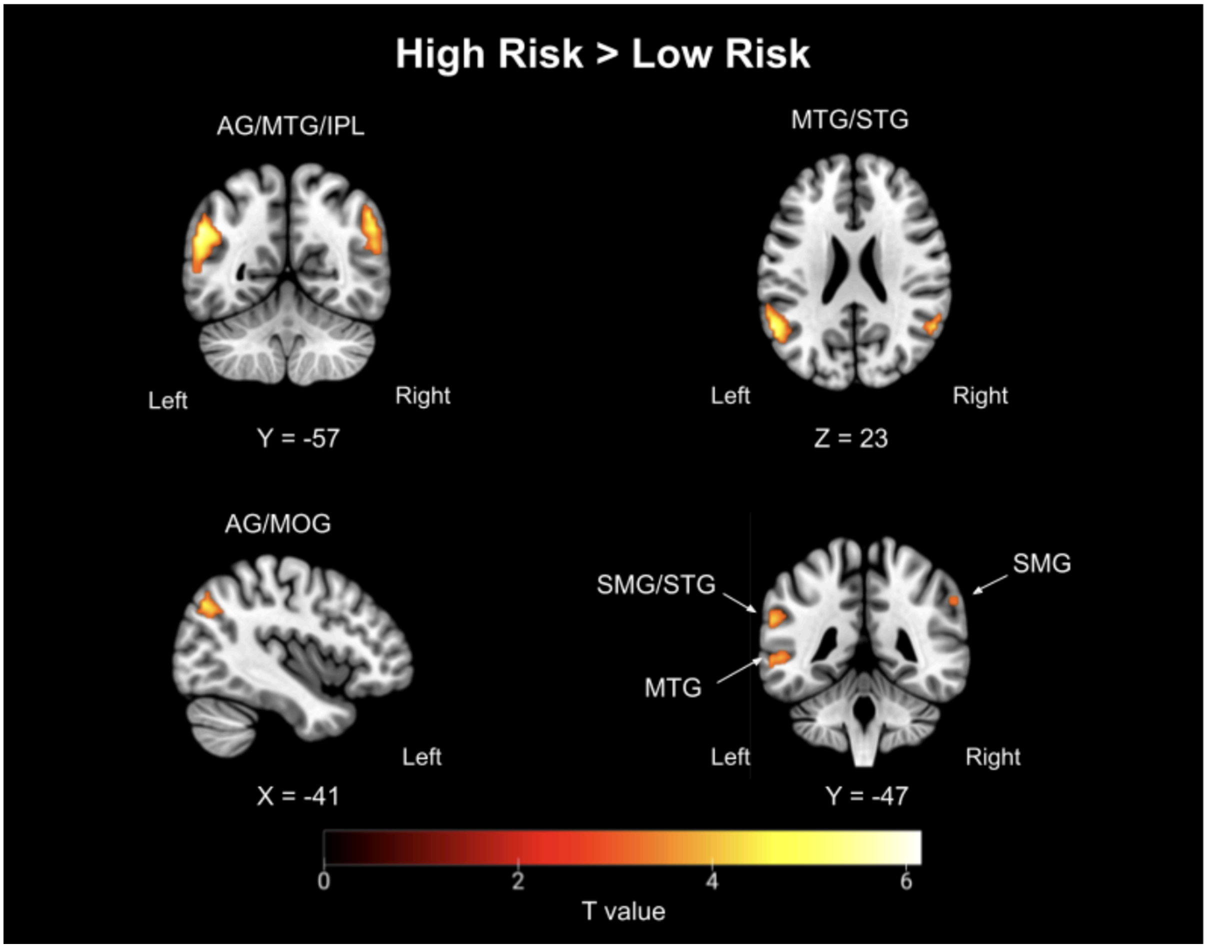


**Figure S3**. Whole brain activation for the High Risk > Low Risk decisions contrast (N = 185; family wise error (FWE) corrected p < .05 at the cluster level, with a cluster forming threshold of p < .001). The T value color bar indicates the strength of activation, ranging from weaker (red) to stronger (yellow). Note: AG, angular gyrus; IPL, inferior parietal lobule; MTG, middle temporal gyrus; STG, superior temporal gyrus, MOG, middle occipital gyrus; SMG, supramarginal gyrus

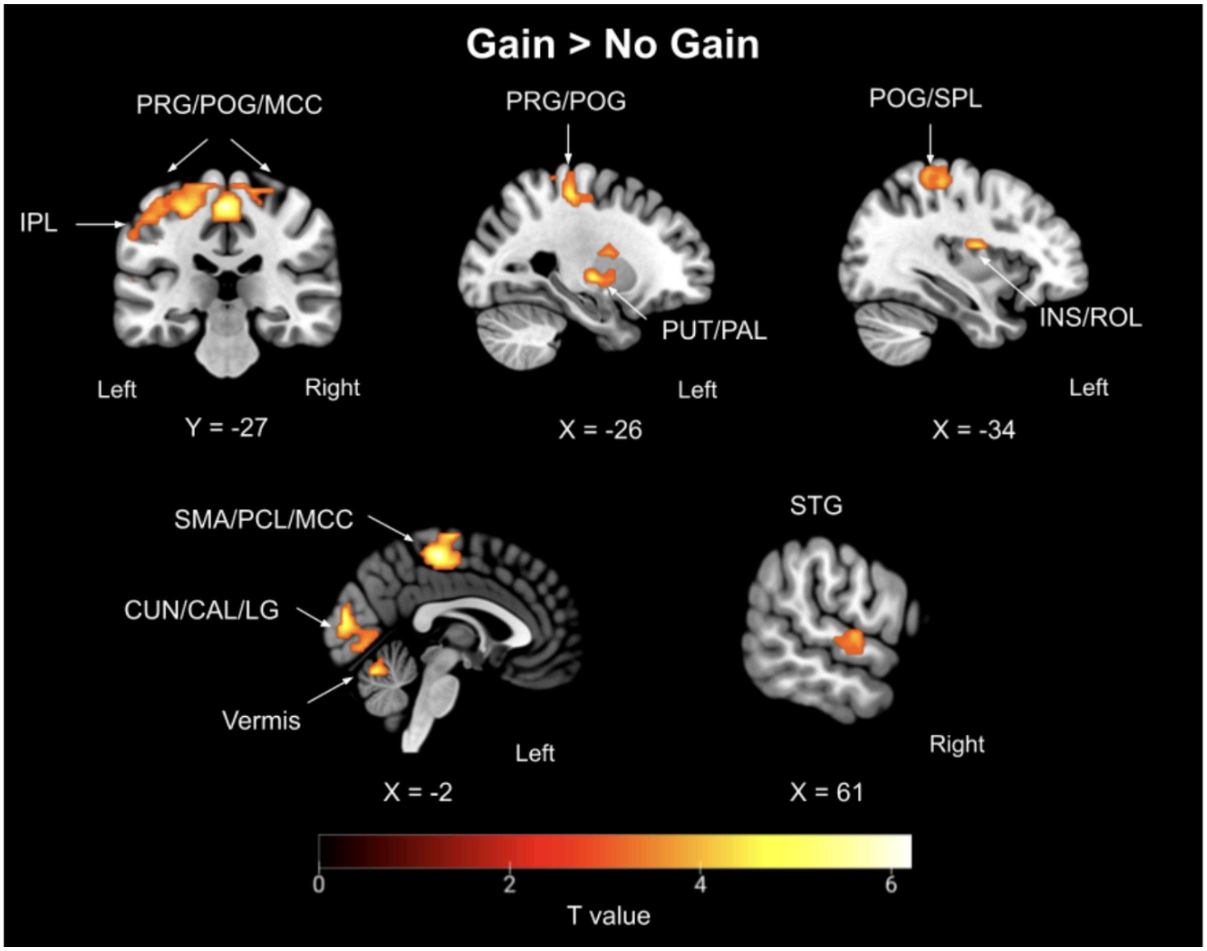


**Figure S4**. Whole brain activation for the Gain > No Gain outcomes contrast (N = 185; family wise error (FWE) corrected p < .05 at the cluster level, with a cluster forming threshold of p < .001). The T value color bar indicates the strength of activation, ranging from weaker (red) to stronger (yellow). Note: PRG, precentral gyrus; POG, postcentral gyrus; IPL, inferior parietal lobule; PUT, putamen; PAL, pallidum; SPL; superior parietal lobule; INS, insula; ROL, rolandic operculum; SMA, supplementary motor area; PCL, paracentral lobule; MCC, middle cingulate & paracingulate gyri; STG, superior temporal gyrus

**Table S2. Clusters with significant activation for High Risk > Low Risk and Gain > No Gain contrasts in PROGRESS-MRI participants with CGT fMRI data (n = 185).**

| Contrast | Brain Region | Hemisphere | MNI Coordinates (x, y, z) | Voxels | T | Cluster-level FWE-corrected *p* |
|---|---|---|---|---|---|---|
| **High Risk > Low Risk** | Angular gyrus, supramarginal gyrus, IPL, STG, MTG | R | 52 -56 43 | 151 | 4.90 | <0.001 |
| | Angular gyrus, supramarginal gyrus, IPL, middle occipital gyrus, MTG, STG | L | -59 -56 25 | 413 | 5.99 | <0.001 |
| | MTG, STG | L | 61 -29 -2 | 80 | 5.19 | 0.007 |
| **Gain > No Gain** | Paracentral lobule, precentral gyrus, postcentral gyrus, SMA, MCC, precuneus, supramarginal gyrus [a], IPL [a], SPL[a] | R/L | -2 -26 58 | 795 | 5.94 | <0.001 |
| | Cuneus, calcarine cortex, lingual gyrus, cerebellum [a] | R/L | -5 -92 19 | 436 | 5.76 | <0.001 |
| | Insula, rolandic operculum, postcentral gyrus, putamen, supramarginal gyrus, STG | L | -35 -8 -16 | 141 | 4.90 | <0.001 |
| | STG, heschl's gyrus | L | 64 -14 1 | 56 | 4.06 | 0.02 |
| | Pallidum, putamen | L | -26 -11 -2 | 46 | 4.80 | 0.04 |

Note: Voxels: number of activated voxels per cluster; T: T-statistic for each cluster; R, right; L, left; MNI, Montreal Neurological Institute; *SMA, supplementary motor area; dACC, dorsal anterior cingulate cortex; IPL, inferior parietal lobule; MTG, middle temporal gyrus; STG, superior temporal gyrus; MCC, middle cingulate & paracingulate gyrus; SPL, superior parietal lobule*

[a]*Activation in left hemisphere only*

**Figure S5. Time-varying associations between early-life metal mixture exposure and reward-related neural activation in functional regions of interest derived from the Gain > No-Gain outcomes contrast.**

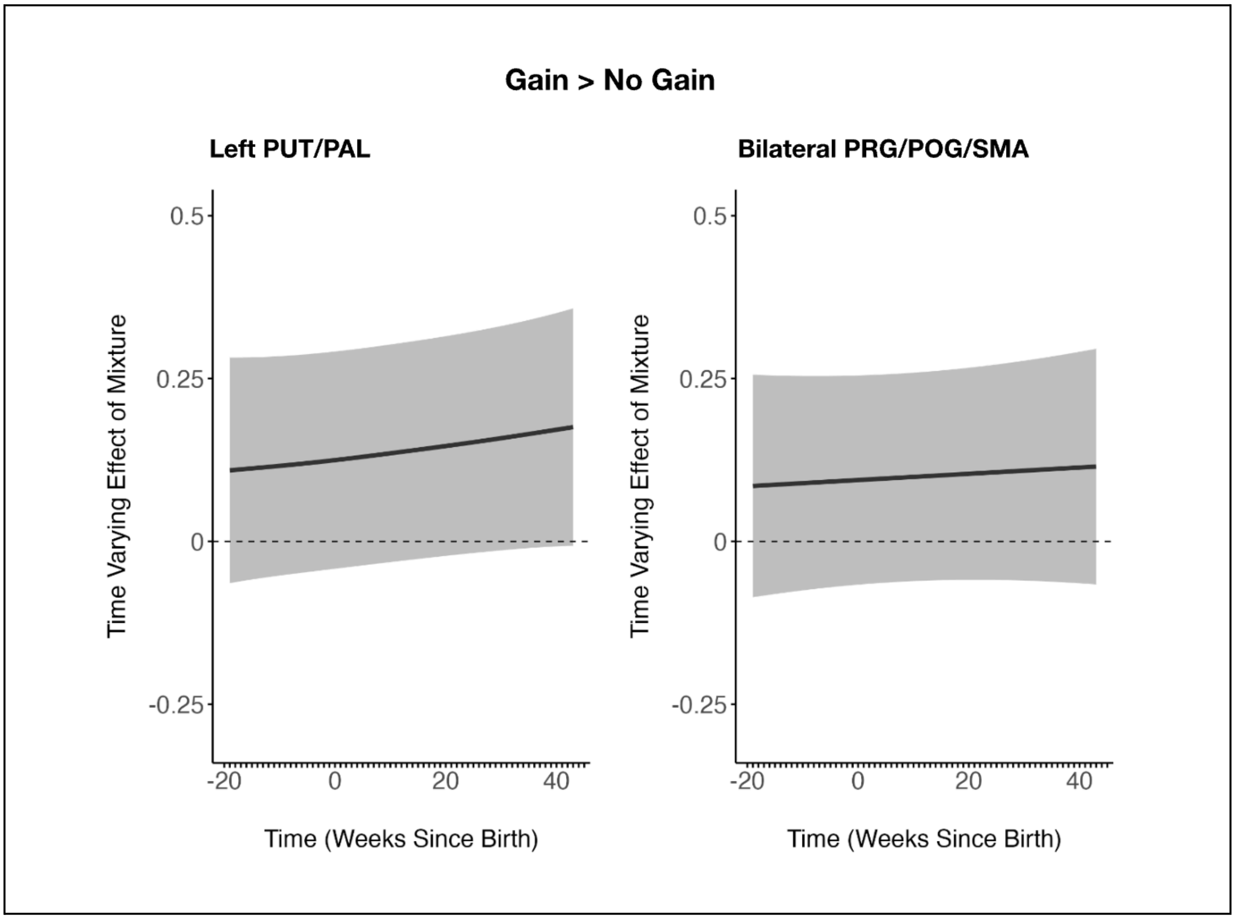


Results from non-significant LWQS models for neural constructs derived from the Gain > No-Gain outcomes contrast (N = 166). Gray shadows display 95% piecewise confidence intervals. The Y-axis represents time-varying correlations between the metal mixture and functional ROIs: left PUT/PAL; bilateral PRG/POG/SMA. The X-axis represents the time since birth, indicating the timing (in weeks) of tooth sampling. Note: PUT/PAL: putamen/pallidum; PRG/POG/SMA: precentral gyrus/postcentral gyrus/supplementary motor area

**Figure S6. Time-varying associations between individual metal exposures and participant-specific reward sensitivity.**

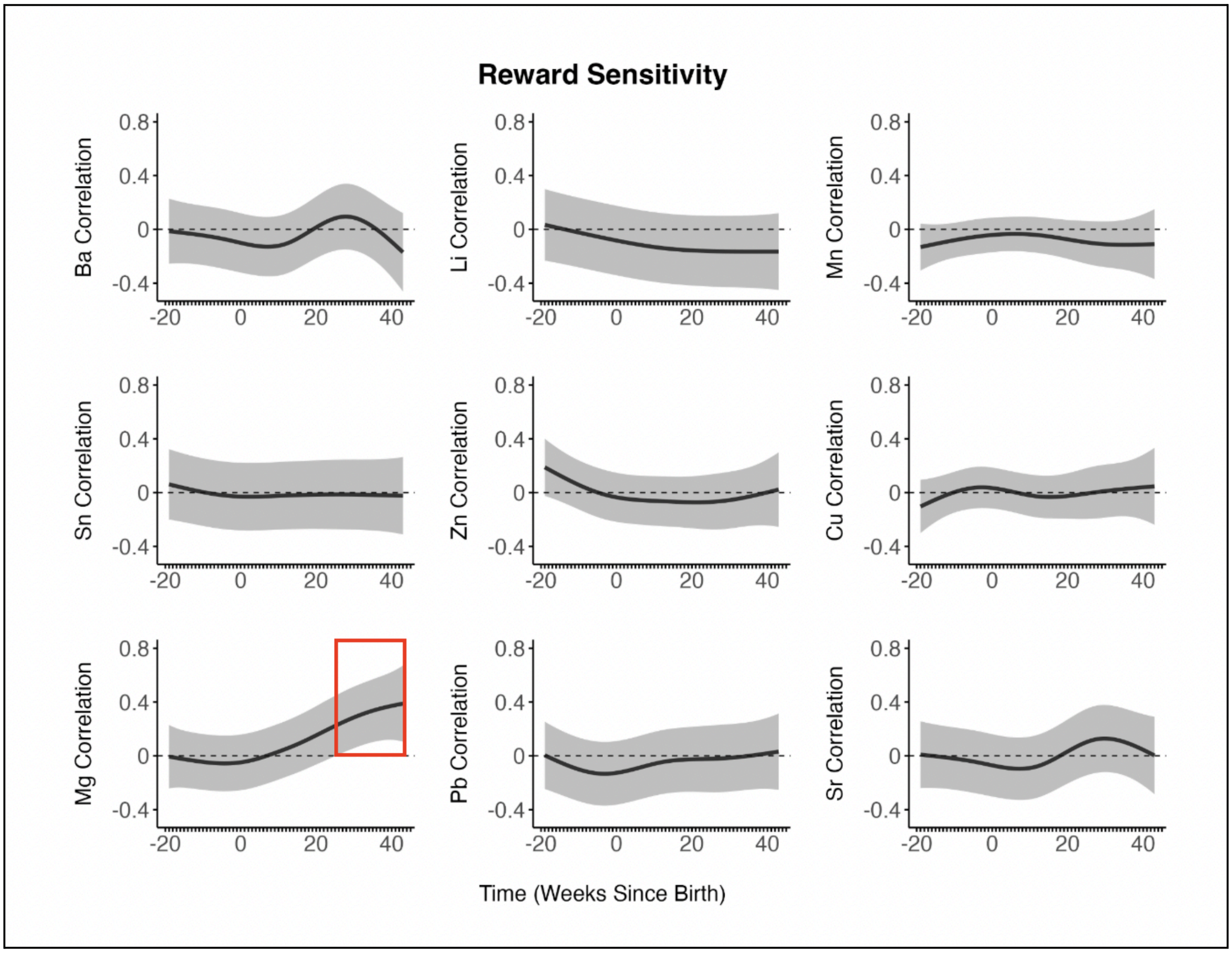


Results from individual reverse DLMs for participant-specific reward sensitivity (N = 189). Gray shadows display 95% piecewise confidence intervals. The Y-axis represents time-varying correlations between single metal concentrations and reward sensitivity. The X-axis represents the time since birth, indicating the timing (in weeks) of tooth sampling. Note: Barium: Ba; Copper: Cu; Lithium: Li; Magnesium: Mg; Manganese: Mn; Lead: Pb; Tin: Sn; Strontium: Sr; Zinc: Zn

**Figure S7. Time-varying associations between individual metal exposures and right angular gyrus/inferior parietal lobule activation derived from the High Risk > Low Risk decisions contrast.**

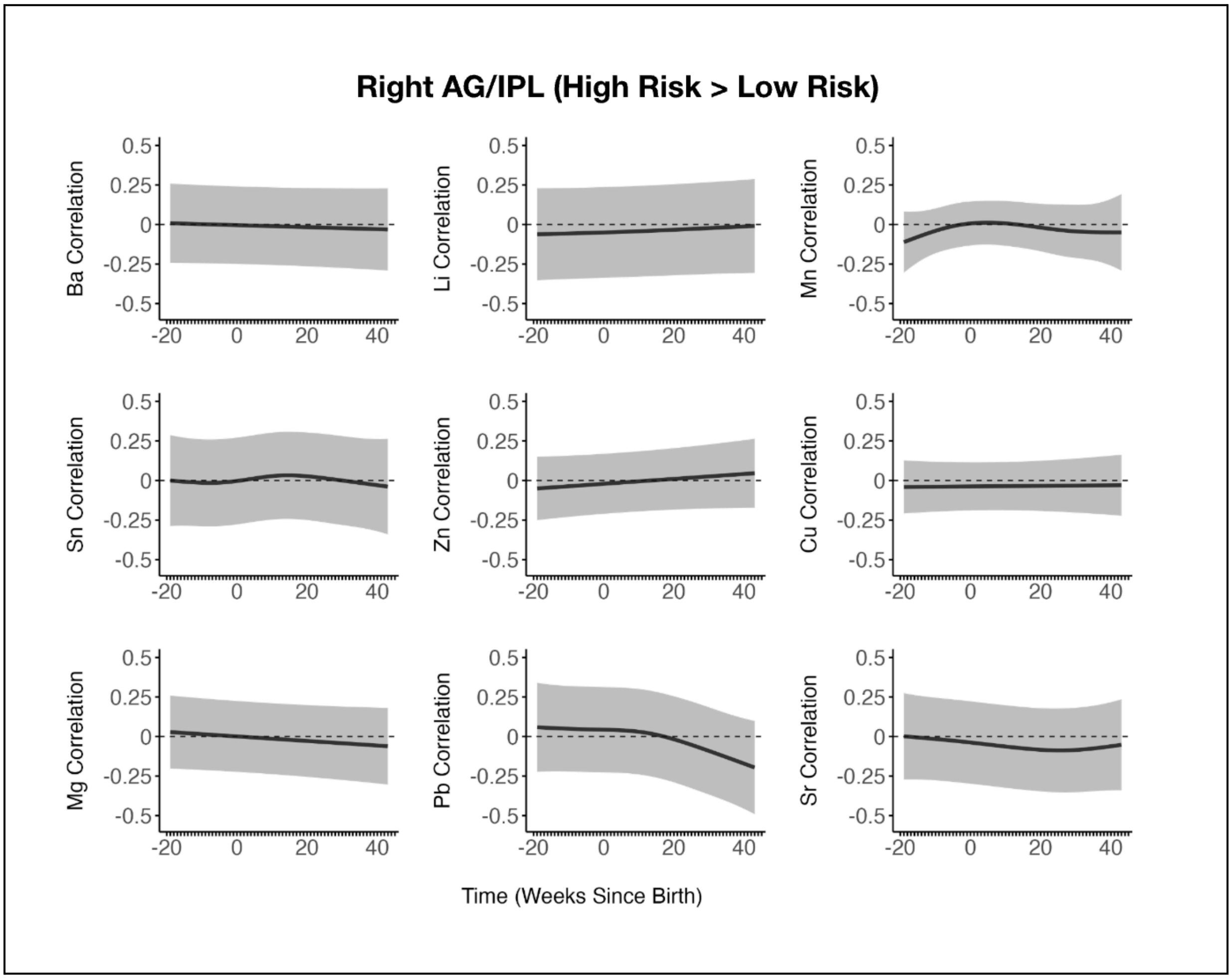


Results from individual reverse DLMs for right angular gyrus/inferior parietal lobule activation derived from the High Risk > Low Risk decisions contrast (N = 166). Gray shadows display 95% piecewise confidence intervals. The Y-axis represents time-varying correlations between single metal concentrations and right angular gyrus/inferior parietal lobule activation. The X-axis represents the time since birth, indicating the timing (in weeks) of tooth sampling. Note: AG: angular gyrus; IPL: inferior parietal lobule; Barium: Ba; Copper: Cu; Lithium: Li; Magnesium: Mg; Manganese: Mn; Lead: Pb; Tin: Sn; Strontium: Sr; Zinc: Zn

**Figure S8. Time-varying associations between individual metal exposures and left insula/rolandic operculum activation derived from the Gain > No Gain outcomes contrast.**

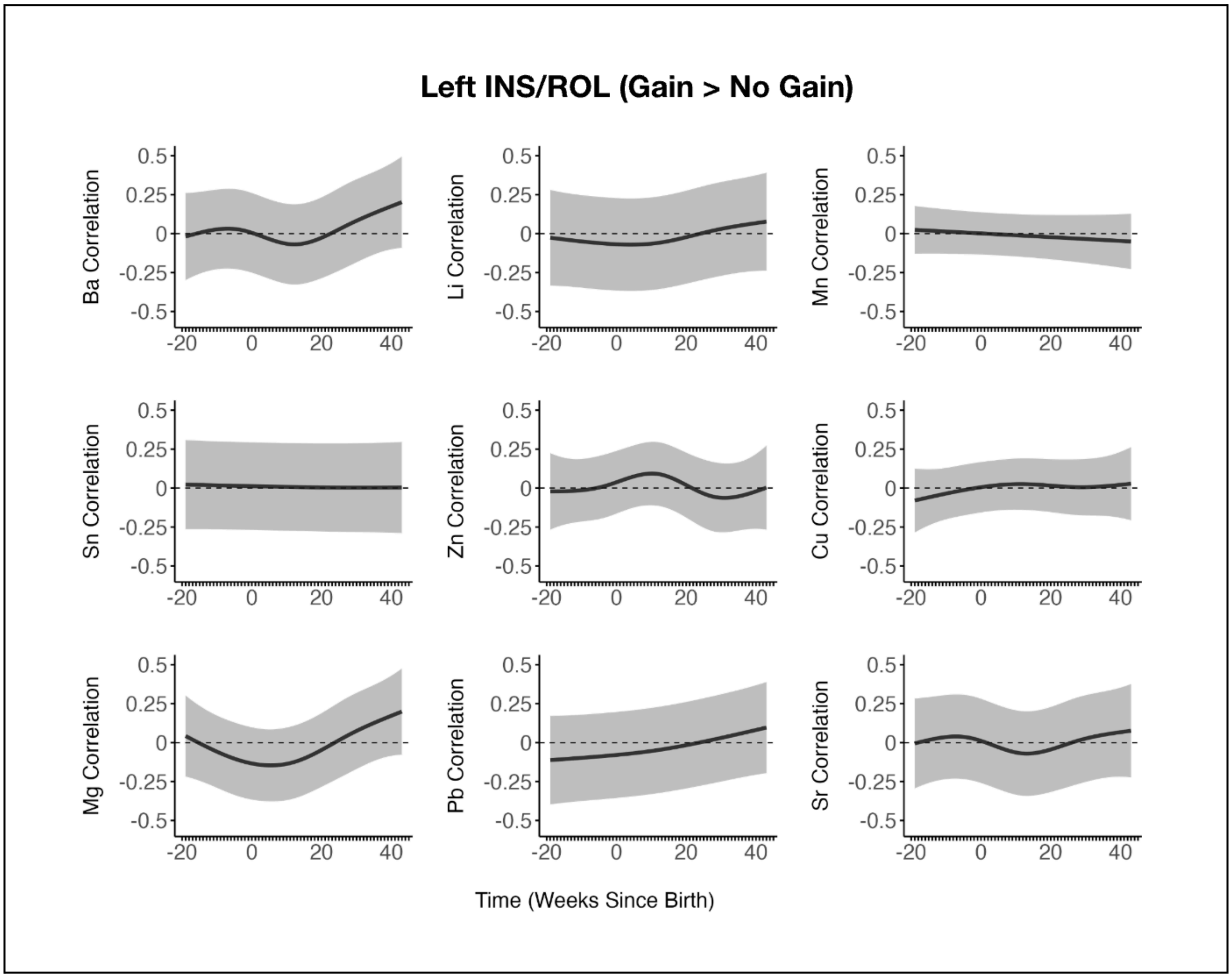


Results from individual reverse DLMs for left insula/rolandic operculum activation derived from the Gain > No Gain outcomes contrast (N = 166). Gray shadows display 95% piecewise confidence intervals. The Y-axis represents time-varying correlations between single metal concentrations and left insula/rolandic operculum activation. The X-axis represents the time since birth, indicating the timing (in weeks) of tooth sampling. Note: INS: insula; ROL: rolandic operculum; Barium: Ba; Copper: Cu; Lithium: Li; Magnesium: Mg; Manganese: Mn; Lead: Pb; Tin: Sn; Strontium: Sr; Zinc: Zn

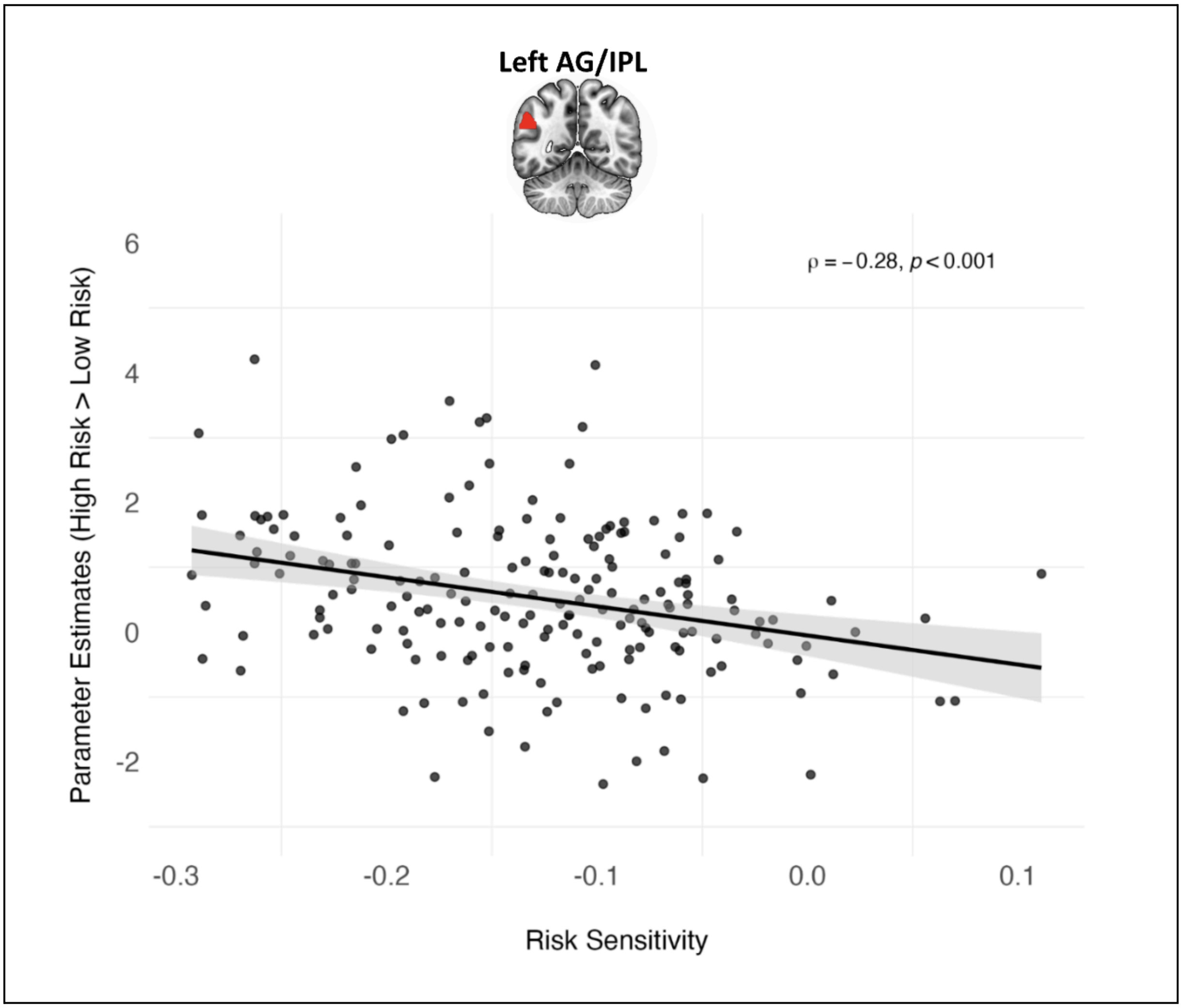


**Figure S9**. Scatterplot showing the correlation between participants' behaviorally estimated risk sensitivity and neural activation during high-risk vs low-risk decisions in the left AG/IPL. Note: AG: angular gyrus; IPL: inferior parietal lobule

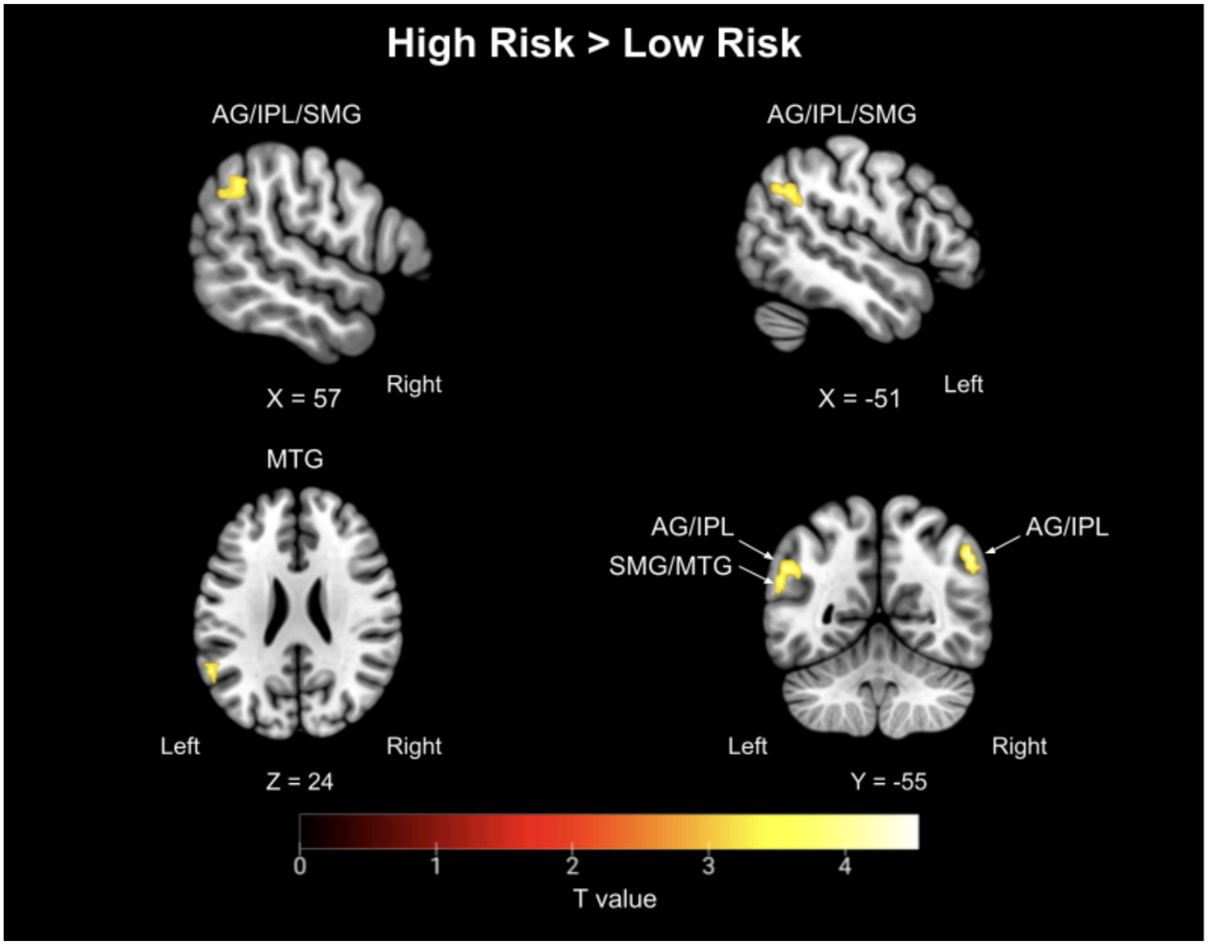


**Figure S10**. Whole brain activation for the High Risk > Low Risk decisions contrast among participants who selected the high risk gamble for $\geq 20\%$ of trials (N = 149; family wise error (FWE) corrected $p < .05$ at the cluster level, with a cluster forming threshold of $p < .001$). The T value color bar indicates the strength of activation, ranging from weaker (red) to stronger (yellow). Note: AG, angular gyrus; IPL, inferior parietal lobule; MTG, middle temporal gyrus; SMG, supramarginal gyrus
